# Health behaviours prior to pregnancy and fertility outcomes: Triangulation of evidence in the Norwegian Mother, Father and Child Cohort Study (MoBa)

**DOI:** 10.1101/2022.03.22.22272772

**Authors:** Robyn E Wootton, Rebecca B Lawn, Maria Magnus, Jorien Treur, Elizabeth Corfield, Pål R. Njølstad, Ole A. Andreassen, Deborah A. Lawlor, Marcus R Munafò, Siri E. Håberg, George Davey Smith, Ted Reichborn-Kjennerud, Per Magnus, Alexandra Havdahl

## Abstract

**Introduction:** Current advice to improve fertility includes reducing alcohol and caffeine consumption, achieving healthy weight-range, and stopping smoking. Advice is informed by observational evidence, which is often biased by confounding.

**Methods:** This study uses data from the Norwegian Mother, Father and Child Cohort Study (MoBa) and the Medical Birth Registry of Norway. First, we analysed associations between health behaviours prior to pregnancy (alcohol and caffeine consumption, body-mass index (BMI) and smoking) and multiple indicators of fertility (including number of children, time to conception, and miscarriage) (*n*=83,128 women, 67,555 men), adjusting for birthyear, education and attention deficit and hyperactive-impulsive (ADHD) traits. Second, we used individual-level Mendelian randomisation (MR) to explore possible causal effects of health behaviours on fertility outcomes (*n*=27,216 women, 26,131 men). Finally, we performed summary-level MR for available outcomes (*n*=91,462-1,232,091) and conducted multi-variable MR to control for education and ADHD liability.

**Results:** In observational analysis, higher BMI and smoking (and to a lesser extent caffeine) were predominantly associated with reduced fertility outcomes. Unexpectedly, higher alcohol consumption was associated with predominantly improved fertility outcomes. There was little evidence from individual-level MR analyses, except smoking and higher BMI were associated with younger age at first birth in women (mean difference in years, per SD increase in genetic score; smoking: -2.65 (95%CI: -3.57, -1.73); BMI: -0.11 (95%CI: -0.16, -0.08)) and men (smoking: -2.82 (95%CI: -4.07, -1.58); BMI: -0.17 (95%CI: -0.23, -0.11)). These results were replicated in the summary-level MR analysis, however effects attenuated after adjusting for education and ADHD liability.

**Conclusions:** Most observational evidence for associations between health behaviours and fertility was not supported by MR analyses, suggesting possible residual confounding. Evidence from MR analyses supported an effect of smoking and higher BMI on younger age of first birth, but multivariable MR suggested this might be explained by underlying liability to ADHD and low educational attainment.

## Introduction

Women struggling to conceive are often advised to engage in healthier lifestyle behaviours, for example, reducing their alcohol and caffeine consumption, achieving a healthy weight-range and quitting smoking (1–3). Reviews of observational evidence support an association between these health behaviours and reduced fertility in women (4), with high alcohol consumption and smoking associated with reduced likelihood of conception (5–9) and high caffeine consumption and obesity associated with increased risk of miscarriage (10,11). Guidelines and epidemiological research on fertility often focus on women (2,12,13) however, it is important to also consider the impact of health behaviours in the partner. Meta-analyses suggest that smoking and alcohol consumption can reduce semen quality (14), and paternal obesity has been associated with reduced likelihood of natural conception (15) and an increased time to conception (16).

The majority of evidence to date exploring health behaviours and fertility is observational, and most studies do not adequately control for confounding (17). For example, health behaviours often co-occur with other health behaviours which might instead affect fertility (e.g. diet, physical activity, sleep). Furthermore, health behaviours and fertility outcomes (such as age at first birth and number of children) share common risk factors including low educational attainment and liability to inattention and hyperactive-impulsive behaviour (traits of attention deficit hyperactivity disorder; ADHD)(18–20). In addition to confounding, it is necessary to account for reverse causation because the stress of being unable to conceive might cause couples to engage in unhealthy behaviours. We extended previous epidemiological studies using Mendelian randomisation (MR), which can reduce bias from residual confounding and reverse causation. MR uses genetic variants (single nucleotide polymorphisms; SNPs) to estimate the causal effect of an exposure on an outcome (21). Causal evidence could improve fertility guidance, helping couples successfully conceive as well as removing any unnecessary stress or guilt around unrelated lifestyle factors. The objective of the current study was to explore the associations between health behaviours and fertility outcomes in both men and women using an MR approach. We used the Norwegian Mother, Father and Child (MoBa) pregnancy cohort, containing around 95,000 mothers and 75,000 partners.

## Methods

### Sample

MoBa is a prospective population-based pregnancy cohort study conducted by the Norwegian Institute of Public Health. Pregnant women were recruited from all over Norway from 1999-2008 (22,23). Women consented to participation in 41% of the pregnancies. The cohort includes 114 500 children, 95 200 mothers and 75 200 partners. The current study was based on version 12 of the quality-assured data files released for research on January 2019. Questionnaires were completed across multiple time points during pregnancy and after birth. The present study used measures from the first questionnaires sent to mothers and fathers, received between 13-17 weeks gestation (hereon referred to as the 15-week questionnaire). The questions related to previous pregnancies, medical history and medication use, occupation, exposures in the workplace and home, lifestyle habits and mental health. In addition to questionnaire data, information on maternal characteristics and pregnancy outcomes of the index pregnancies were available through data linkage to the Medical Birth Registry Norway (MBRN). MBRN also provided data on age at first birth and total number of children up to 2018, including (but not limited to) the MoBa index pregnancy. The establishment of MoBa and initial data collection was based on a license from the Norwegian Data Protection Agency and approval from The Regional Committees for Medical and Health Research Ethics. The MoBa cohort is now based on regulations related to the Norwegian Health Registry Act. The current study was approved by The Regional Committees for Medical and Health Research Ethics (2016/1702).

After restricting to those with available exposure and outcome data, a maximum of 83,128 women and 67,555 men were included in multivariable regression analysis. After further restricting to those individuals with quality-controlled genotype data (see details below), a maximum of 27,216 women and 26,131 men were available for individual-level MR analysis. More details of participant exclusion are given in Supplementary Figure S1.

### Health behaviours exposure data

For all health behaviours, we used data from the 15-week questionnaire in pregnancy. Women were asked to report their behaviour 3 months prior to the index pregnancy. Fathers were asked to report their behaviour 6 months prior to the index pregnancy.

#### Alcohol consumption

Frequency of alcohol consumption was self-reported on a 7-point scale (never, less than once a month, 1-3 times a month, once per week, 2-3 times per week, 4-5 times per week, 6-7 times per week). Women and men were also asked about binge drinking behaviour: “How often did you drink 5 units or more on one occasion?”. Responses were on a 5-point scale (never, less than once a month, 1-3 times a month, once a week, several times per week).

#### Caffeine consumption

Caffeine consumption in women was calculated from self-reported daily beverage consumption, where one cup contained 125ml. Caffeine (mg) weights per cup were taken from a previous study of caffeine consumption in MoBa (24). We excluded outliers if they consumed more than 3.5 litres of any particular drink in a day (28 cups) or if their total caffeine consumption per day was greater than 1000mg (25). Values were log transformed to adjust for skewness. In men, beverage consumption was instead measured categorically over a typical week. Responses were on a 5-point scale (seldom/never, 1-6 cups a week, 1 cup a day, 2-3 cups a day, 4+ cups a day). Unlike the questionnaire administered to women, men were not directed as to the volume of the cup, therefore we followed previous calculations (24) and assumed that a cup was 125ml for coffee (apart from espresso where we assumed a standard single is 30ml) and 250ml for tea or fizzy drink. Again consumption was weighted by caffeine (mg) (24) and divided by 7 to estimate mg per day.

#### Smoking behaviour

Smoking initiation was self-reported: “Have you ever smoked?” where yes was classed as ever smoking and no was classed as never smoking. Smoking heaviness was self-reported amongst current smokers as the average number of cigarettes smoked per day prior to pregnancy. We excluded current smokers who reported smoking no cigarettes.

#### Body mass index (BMI)

Height and weight were self-reported. We used pre-pregnancy values for women and current (15-weeks) values for men. Women were also asked to report their partner’s height and weight. These reports were highly correlated with partners’ self-report (r=0.98 for height and r=0.95 for weight). Therefore, we used the woman’s report of their partner’s height and weight when the partner’s own report was unavailable. We excluded outliers for women at height <117 or >196 cm and weight <38 or >150 kg and for men at height <136 or >220 cm and weight <50 or >200 kg as done previously in MoBa (16). From these height and weight measures we calculated BMI as weight (kg)/height (m^2^). We note BMI is not a health behaviour itself but is a biomarker which can crudely proxy for healthy behaviours.

### Fertility outcome data

All fertility outcomes were self-reported in the 15-week questionnaire by the women apart from age at first birth and total number of children, which were obtained from the Medical Birth Registry of Norway (MBRN) (version 12, last update November 2018).

#### Age at first birth

Age at the time of first recorded child’s delivery obtained from MBRN (not limited to births recorded in MoBa).

#### Number of children

The total number of children born to women and men up to November 2018 obtained from MBRN (including but not limited to births recorded in MoBa). Totals of more than 6 children were grouped into a 6+ category to adjust for skewness.

#### Time to conception

Based upon women’s self-report from the 15-week questionnaire referring to the index pregnancy. If women reported planning their pregnancy, they were asked: “For how many months did you have regular intercourse without contraception before you became pregnant?”. Options were less than one month, 1-2 months or 3+ months. If it took more than 3 months, then they were asked to state the number of months. We combined anyone taking 12 or more months to conceive into one group to reduce skewness and treated as a continuous variable. We have used this woman-reported variable as an outcome in both women and men under the assumption that couples were conceiving together. In our primary analysis, if the couple were not trying to conceive then they were set to missing. However, given differences between couples who planned pregnancy and those who did not, we also conducted a sensitivity analysis where non-planners were included and assigned them the median time to conception from the planning group (2 months) (Supplementary Note 2). We also conducted a sensitivity analysis using dichotomised variables as time to conception was not measured continuously (Supplementary Note 3).

#### Infertility treatment

Women self-reported in the 15-week questionnaire: “Have you ever been treated for infertility?”. Responses were binary *yes* or *no*. We did not use this variable as an outcome in the fathers, as this question did not specify whether the infertility treatment was for the index pregnancy.

#### Miscarriage

Women self-reported (in the 15-week questionnaire) ever having had a miscarriage, defined as any of their previous pregnancies ending in spontaneous abortion at or before the 20^th^ week of pregnancy.

As a secondary outcome, we also explored the impact of health behaviours on frequency of sexual intercourse (see Supplementary Note 4).

### Genotype data

Blood samples were obtained from both parents during pregnancy and from children (umbilical cord) at birth (26). For the current study we used MoBa Genetics genotype data release 1.0 (https://github.com/folkehelseinstituttet/mobagen/wiki/MoBaGenetics1.0). This release has genotype data available for 99,137 individuals (mostly family trios). Details of the genotyping and QC procedures are available in Supplementary Note 1 (27) and Supplementary Figure S1. After quality control and relatedness checks, the remaining samples contained 28,929 women and 27,723 men.

### Genetic score construction

For individual-level MR, our genetic instruments were individual-level genetic scores constructed using PRSice (28) and genome-wide significant variants were possible. SNPs were clumped to ensure independence (r2 < 0.01, clumping window < 1000kb) and weighted by effect sizes from discovery GWAS detailed below (selected for being the largest samples using individuals of European ancestry and not containing the MoBa cohort). Prior to analysis, we checked that each genetic score explained significant variance in the exposure. This is presented in Supplementary Table S1 along with number of SNPs passing QC in the MoBa cohort.

### GWAS Summary Statistics for health behaviours

The following GWAS summary statistics were used to construct genetic scores for individual-level MR, and individual SNP effects sizes were used in summary-level MR.

#### Alcohol consumption

We used two genetic instruments for alcohol consumption: 1) alcohol consumption frequency and 2) binge drinking. Alcohol consumption frequency was measured as average number of drinks per week aggregated across types of alcoholic beverage. The GWAS identified 99 conditionally independent genome-wide significant SNPs in a sample of 941,280 individuals, explaining 2.5% of the variance (29). Binge drinking in the UK Biobank was defined as “How often do you have six or more drinks on one occasion?”, where a drink was defined as a unit of alcohol. The GWAS was conducted by the Neale Lab (http://www.nealelab.is/uk-biobank – round 2, August 2018), and variance explained in an independent sample was not reported. After restricting to independent variants, 4 genome-wide significant SNPs remained. Given the small number of genome-wide significant SNPs, we also used a relaxed p-value threshold of p<5×10^−6^, for which 12 SNPs were available.

#### Caffeine consumption

Caffeine consumption was measured as number of cups of coffee per day. The GWAS identified 6 independent genome-wide significant SNPs in a sample of 91,462 coffee drinkers of European ancestry (30). Genome-wide significant SNPs explained 1.3% of the variance in coffee consumption. Given the small number of genome-wide significant SNPs, we also used a relaxed p-value threshold of p<5×10^−6^, for which 39 SNPs were available.

#### Smoking behaviour

We used two genetic instruments for smoking behaviour: 1) smoking initiation and 2) smoking heaviness. Smoking initiation was defined as ever v. never regularly smoking (more than 100 cigarettes ever or having ever been a daily smoker). The GWAS of smoking initiation identified 378 conditionally independent genome-wide significant SNPs, in a sample of 1,232,091 individuals, which explained 4% of the variance (29). Smoking heaviness was defined as average number of cigarettes smoked per day. The GWAS of smoking heaviness identified 55 conditionally independent genome-wide significant SNPs in a sample of 337,334 ever smokers, which explained 4% of the variance (29). We also conducted a single-SNP analysis of rs16969968 genotype from the CHRNA5 gene, known to reduce nicotine aversion and consequently increase cigarettes smoked per day (31) (see Supplementary Note 5).

#### Body mass index (BMI)

The most recent GWAS of adult BMI identified 941 independent genome-wide significant SNPs in a sample of 681,275 individuals of European ancestry, which explain 6% of the variance in BMI (32).

### Statistical analysis

All analyses were conducted in R version 4.0.3 (33) and performed separately for women and men. We corrected for multiple testing using a Bonferroni correction of 0.05/48 tests (6 exposures and 8 outcomes) which resulted in an adjusted p-value of p<0.001.

#### Stage 1. Multivariable regression analyses

We first explored the association between each of the fertility outcomes using correlation for continuous traits, chi-squared tests for binary traits and independent t-tests when one trait was continuous and the other binary. Second, we conducted linear regressions for each of the continuous fertility outcomes and logistic regressions for each of the binary fertility outcomes to see if we observed the expected association between health behaviours and fertility. Results are presented unadjusted and adjusted for birth year and educational attainment (at around 15-weeks pregnancy). An additional adjustment for ADHD traits was included as a sensitivity analysis to proxy for ADHD liability. ADHD traits were measured when the index child was age 3 years using the Adult ADHD Self-Report Scale (34).

#### Stage 2. Individual-level Mendelian randomisation

MR can be implemented as an instrumental variable analysis using genetic variants to proxy for an exposure. It can be used to estimate a causal effect of the exposure (health behaviours) on the outcome (fertility) providing certain assumptions are satisfied (21,35). The three core assumptions for valid causal inference are: 1) relevance – the genetic instrument must be robustly associated with the exposure, 2) independence – there should be no confounding between the genetic instrument and outcome, and 3) exclusion-restriction – the genetic instrument must only be associated with the outcome via the exposure. Additionally, for results to generalise to other populations, there must be no “defiers” – individuals whose exposure is opposite to their genetic predisposition (35). Our individual-level MR analysis used individual-level genetic scores (with weights from external independent GWAS) in an instrumental variable regression controlling for age, genotyping chip and top 10 principal components of population structure. The genetic scores are first regressed on the exposure, and then predicted values are regressed onto the outcome. Analyses were performed using the *ivreg* command from the Applied Econometrics with R (*AER)* package for R. For continuous outcomes, betas are the mean difference per SD increase in the genetic score. When outcomes are binary, estimates approximate the risk difference per SD increase in genetic score.

#### Sensitivity analyses

We checked for evidence of assortative mating by using the woman’s genetic score to predict the men’s health behaviours and vice versa and by estimating the correlation between the genetic score of women and men. We checked for evidence of possible pleiotropy by testing whether each of the genetic scores predicted any known confounders of the exposure-outcome association (e.g., other health behaviours, income, age) and compared these estimates with the estimated association between observed exposures on confounders. Pleiotropy occurs when one genetic variant influences multiple phenotypes. If these other phenotypes are not on the causal pathway from exposure to outcome (horizontal pleiotropy) then the independence and exclusion-restriction assumptions could be violated, and genetic variants are invalid. Where there was evidence for a causal effect in the individual-level MR analysis, we followed up with additional summary-level MR sensitivity analyses (MR Egger (36), weighted median (37) and weighted mode (38)) which make different assumptions about the nature of pleiotropy. A consistent direction of effect across the different MR sensitivity analyses gives us more confidence that the effects are not due to pleiotropy. We also calculated the MR Egger intercept. If the intercept is significantly different from zero, then this suggests significant directional pleiotropy may be biasing the estimate. To conduct these summary-level sensitivity MR analyses, we generated SNP-outcome association results for the MoBa cohort using the *assoc* command for plink (version 2) (39).

#### Stage 3. Summary-level Mendelian randomisation

The second MR method used was summary-level MR (i.e. two-sample MR) which uses summary statistics from published GWAS (40). Here we don’t have an effect estimate for each individual but instead an effect size for each SNP from the discovery GWAS and an effect size for that SNP in an outcome GWAS. The ratio of these two effect sizes can be meta-analysed across multiple SNPs to give us an estimate of the causal effect. This is our primary analysis known as the inverse-variance weighted estimate. As sensitivity analyses, again we conducted MR Egger (36), weighted median (37) and weighted mode (38), which each make orthogonal assumptions about the nature of pleiotropy.

Independent summary GWAS data was available for three of the outcomes: age at first birth (N= 170,498), number of children (N=333,628) and miscarriage (N= 78,700), self-reported in the UK Biobank (41). We used outcome GWAS from the UK Biobank only to prevent sample overlap with our health behaviour exposure GWAS. For age at first birth and miscarriage, we used GWAS summary statistics from the MRC IEU Open GWAS project (42). The GWAS for age at first birth used the self-reported question “*How old were you when you had your first child?*” (field 2754) asked only to women who had previously indicated that they had given birth to at least one child. The GWAS for miscarriage used the item “*How many spontaneous miscarriages have you had?*” (field 3839) which was only asked to women who had previously indicated that they had ever had a miscarriage, abortion or stillbirth (field 2774). Finally, the GWAS for number of children combined items “*How many children have you fathered?*” (field 2405) in men and “*How many children have you given birth to? (Please include live births only)*” (field 2734) in women (43). Age at first birth and miscarriage GWAS are in SD units and units for number of children are number of children.

For each of the exposures, we used the same health behaviour GWAS as for the individual-level MR, with the exception of BMI. Here we used an earlier GWAS (44) that did not contain the UK Biobank to avoid sample overlap which can bias estimates (45). For smoking initiation and alcohol consumption, we used SNP-exposure estimates from summary statistics with the UK Biobank and 23andMe removed. Smoking heaviness could not be used an as exposure as the fertility outcome GWASs contain both smokers and non-smokers. Binge drinking could not be included as this GWAS was also conducted in the UK Biobank.

#### Sensitivity analyses

The Cochran’s Q test of heterogeneity was conducted to estimate possible pleiotropy and the MR Egger intercept was estimated to test for bias from directional horizontal pleiotropy. The regression dilution I^2^_GX_ was calculated to assess the suitability of the MR Egger effect estimate and a SIMEX correction applied where necessary (46). Steiger filtering was conducted to check for evidence of reverse causation (47). The mean F statistic was calculated as an indicator of instrument strength, where F < 10 is considered to indicate a weak instrument. Where there was evidence for a causal effect, we conducted multivariable MR analysis (48) to explore possible pleiotropy via education and ADHD liability (see Supplementary Materials for details and Table S20).

## Results

Average health behaviours and fertility outcomes were relatively consistent between the full MoBa sample and the genotyped sub-sample (Table 1). Prevalence of smoking, high alcohol consumption, high caffeine consumption and high BMI were greater in men than women prior to pregnancy and men were older on average than women. Fertility outcomes did not differ between women and men, with the exception of age at first birth which was older in men. Results for frequency of sexual intercourse, and dichotomised time to conception are given in Supplementary Notes 3 and 4. Associations between the different fertility outcomes found that a younger age at first birth was associated with having more children in total, shorter time to conception, being less likely to miscarry and less likely to have infertility treatment (Supplementary Table S2). In the following section we highlight results that passed Bonferroni correction.

**Table 1.** Descriptive statistics comparing exposure and outcome data across mothers and fathers.

|  | Mothers |  |  |  | Fathers |  |  |  |
| --- | --- | --- | --- | --- | --- | --- | --- | --- |
|  | Full Sample |  | Genotyped Sample |  | Full Sample |  | Genotyped Sample |  |
|  | N | Mean (SD)/% | N | Mean (SD)/% | N | Mean (SD)/% | N | Mean (SD)/% |
| **Age (years)** | 94,418 | 30.61 (20.66) | 28,849 | 30.34 (15.55) | 74,409 | 33.05 (18.73) | 27,598 | 32.90 (17.44) |
| **BMI (kg/m2)** | 83,191 | 24.05 (4.30) | 27,272 | 24.08 (4.25) | 67,679* | 25.88 (3.34) | 26,161* | 25.93 (3.33) |
| **Alcohol Consumption** |  |  |  |  |  |  |  |  |
| *Total* | 79,562 | - | 26,819 | - | 64,023 | - | 24,775 | - |
| *Never* | 5,690 | 7.15% | 1,885 | 7.03% | 1,441 | 2.25% | 473 | 1.91% |
| *Less than once a month* | 23,618 | 29.69% | 7,848 | 29.26% | 11,203 | 17.50% | 4,220 | 17.03% |
| *1-3 times a month* | 28,575 | 35.92% | 9,804 | 36.56% | 23,497 | 36.70% | 9,304 | 37.55% |
| *Once per week* | 14,263 | 17.93% | 4,811 | 17.94% | 15,869 | 24.79% | 6,250 | 25.23% |
| *2-3 times per week* | 6,527 | 8.20% | 2,188 | 8.16% | 10,168 | 15.88% | 3,896 | 15.73% |
| *4-5 times per week* | 724 | 0.91% | 221 | 0.82% | 1,488 | 2.32% | 521 | 2.10% |
| *6-7 times per week* | 165 | 0.21% | 62 | 0.23% | 357 | 0.56% | 111 | 0.45% |
| **Binge Drinking** |  |  |  |  |  |  |  |  |
| *Total* | 78,615 | - | 26,546 | - | 28,699 | - | 10,761 | - |
| *Never* | 26,229 | 33.36% | 8,707 | 32.80% | 3,001 | 10.46% | 1,058 | 9.83% |
| *Less than once a month* | 31,002 | 39.44% | 10,647 | 40.11% | 11,726 | 40.86% | 4,575 | 42.51% |
| *1-3 times a month* | 16,428 | 20.90% | 5,606 | 21.12% | 9,534 | 33.22% | 3,545 | 32.94% |
| *Once a week* | 4,313 | 5.49% | 1,411 | 5.32% | 3,732 | 13.00% | 1,348 | 12.53% |
| *Several times per week* | 643 | 0.82% | 175 | 0.66% | 706 | 2.46% | 235 | 2.18% |
| **Smoking Initiation** |  |  |  |  |  |  |  |  |
| *Total* | 82,824 | - | 27,601 | - | 65,670 | - | 25,324 | - |
| *Never smokers* | 41,326 | 49.90% | 13,897 | 50.35% | 30,993 | 47.20% | 12,124 | 47.88% |
| *Ever smokers* | 41,498 | 50.10% | 13,704 | 49.65% | 34,677 | 52.80% | 13,200 | 52.12% |
| **Smoking Heaviness (cigarettes per day)** | 16,074 | 11.41 (5.94) | 4,961 | 11.38 (5.85) | 12,606 | 13.49 (6.39) | 4,721 | 13.42 (6.17) |
| **Caffeine Consumption (mg per day)** | 75,172 | 141.60 (138.36) | 24,512 | 139.40 (136.63) | 27,501 | 125.40 (85.60) | 10,326 | 125.79 (84.99) |
| **Age at First Birth (years)** | 94,740 | 27.15 (4.65) | 28,871 | 27.32 (4.42) | 74,681 | 29.61 (4.99) | 27,617 | 29.63 (4.82) |
| **Number of children (N children)** | 94,740 | 2.55 (0.95) | 28,871 | 2.56 (0.89) | 74,681 | 2.51 (0.91) | 27,617 | 2.52 (0.88) |
| **Time to Conception (months)** | 63,068 | 4.90 (7.94) | 21,749 | 4.63 (7.39) | 53,516∇ | 4.86 (7.84) | 21,146∇ | 4.64 (7.40) |
| **Infertility treatment** |  |  |  |  |  |  |  |  |
| *Total* | 83,876 | - | 27,616 | - | - | - | - | - |
| *Never* | 76,195 | 90.84% | 25,253 | 91.44% | - | - | - | - |
| *Ever* | 7,681 | 9.16% | 2,363 | 8.56% | - | - | - | - |
| **Miscarriage** |  |  |  |  |  |  |  |  |
| *Total* | 55,696 | - | 17,896 | - | - | - | - | - |
| *Never* | 39,114 | 70.23% | 12,617 | 70.50% | - | - | - | - |
| *Ever* | 16,582 | 29.77% | 5,279 | 29.50% | - | - | - | - |

Note. \* = supplemented with mother's report when father's report was unavailable. ∇ = Father variable reported by the mother.

### Stage 1. Multivariable Regression Associations

The results of the observed associations between the health behaviours and fertility outcomes (adjusted for birth year and education) are given in Figure 1 (Tables S3 and S4).

**Figure 1.**
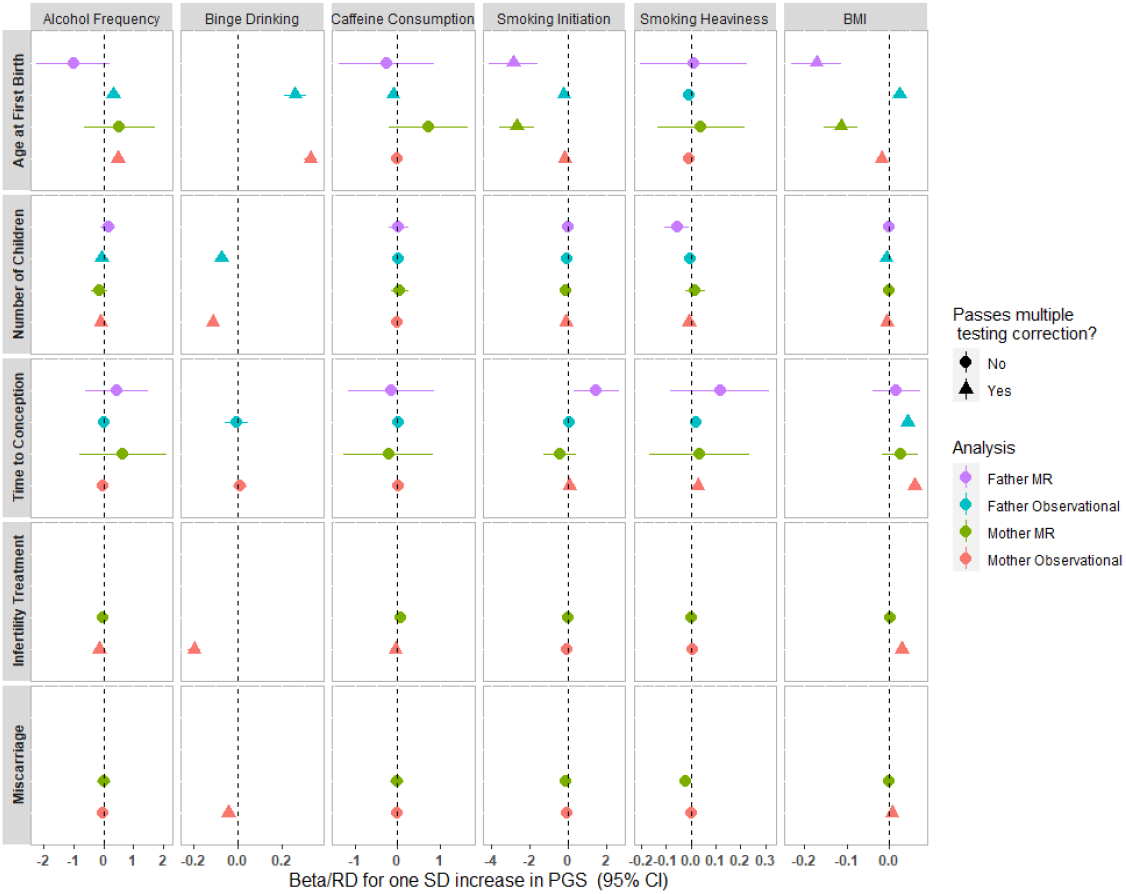
The association between health behaviours and fertility outcomes comparing mothers and fathers, and comparing multivariable regression associations with estimates of causal effects from Mendelian randomisation. For continuous outcomes, units are betas. For binary outcomes, regression units are on the log odds scale and MR units are risk differences.

#### Alcohol Consumption

Units are per category increase in self-reported alcohol consumption or binge drinking. In women’s adjusted analyses, greater frequency of alcohol consumption and binge drinking were both associated with having fewer children (alcohol frequency: -0.090 children, 95% CI: -0.096, -0.085; binge drinking: -0.111 children, 95% CI: -0.118, -0.104) and an older age at first birth (alcohol frequency: 0.492 years, 95% CI: 0.468, 0.516; binge drinking: 0.333 years, 95% CI: 0.304, 0.362). However, those who consumed more alcohol were less likely to have had infertility treatment (alcohol frequency: OR = 0.867, 95% CI: 0.847, 0.888; binge drinking: OR = 0.822, 95% CI: 0.798, 0.847). Binge drinking (but not alcohol frequency) was associated with decreased odds of miscarriage (OR = 0.959, 95% CI: 0.938, 0.981). Neither alcohol frequency nor binge drinking were associated with time to conception after correction for multiple testing. The pattern of results was consistent for outcomes available in men.

#### Caffeine consumption

Units are per unit increase in log transformed mg of caffeine per day. In men, higher caffeine consumption was associated with a younger age at first birth (−0.100 years, 95% CI: -0.139, -0.061) and being less likely to have fertility treatment (OR = 0.951, 95% CI: 0.931, 0.972). Caffeine consumption was not associated with any of the other fertility outcomes in men nor with any fertility outcomes in women correction for multiple testing.

#### Smoking behaviour

Ever smoking was associated with having fewer children in women (−0.101 children, 95% CI: -0.114, -0.089), a younger age at first birth in both women and men (women: - 0.155 years, 95% CI: -0.208, -0.103; men: -0.218 years, 95% CI: -0.284, -0.151), and an increased time to conception in women (0.095 months, 95% CI: 0.038, 0.151). Smoking heaviness was also associated with having fewer children in women (−0.008 children, 95% CI: -0.010, -0.005) and longer time to conception (women: 0.029 months, 95% CI: 0.016, 0.042; men: 0.021 months, 95% CI: 0.009, 0.034).

#### Body mass index

Units are per kg/m^2^ increase in BMI. Higher BMI was associated with worse outcomes across all indicators for fertility in both sexes, including having fewer children (women: -0.004 children, 95% CI: -0.005, -0.002; men: -0.006 children, 95% CI: -0.008, -0.004) and taking longer to conceive (women: 0.061 months, 95% CI: 0.055, 0.068; men: 0.045 months, 95% CI: 0.035, 0.054). For women, having a higher BMI was also associated with being more likely to have fertility treatment (OR: 1.032, 95% CI:1.026, 1.037) and being more likely to have a miscarriage (OR: 1.008, 95% CI: 1.004, 1.013). In men, higher BMI was associated with an older age at first birth (0.026 years, 95% CI: 0.016, 0.035). The only exception to this pattern of worsened fertility was an association between higher BMI in women and younger age of first birth (−0.017 years, 95% CI: -0.023, -0.011).

These results were relatively consistent, but less precise to those observed when restricting the analysis to the genotyped sample only and after adjustment for ADHD traits (Supplementary Tables S5 and S6).

### Stage 2. Individual-level Mendelian randomisation

Genetic scores were associated with the exposures in MoBa with the exception of binge drinking, which was therefore not used in further analyses (Table S1). Results of the individual level MR are presented in Figure 1 (and Tables S7 for women and S8 for men). All genetic scores were standardised, so that the results are per standard deviation increase in genetic score for the exposure. Genetic liability for higher BMI was associated with a younger age of first birth in both women (−0.113 years, 95% CI: -0.153, -0.073) and men (−0.171 years, 95% CI: -0.232, -0.111). Genetic liability for smoking initiation was also strongly associated with a younger age of first birth in both women (−2.647 years, 95% CI: -3.565, -1.730) and men (−2.824 years, 95% CI: - 4.072, -1.575). However, stratified by smoking status, rs16969968 genotype was only associated with age at first birth in never smokers, suggesting possible pleiotropy (Supplementary Note 5 and Figure S2). We found no robust evidence for association between any of the other exposure and outcomes using individual-level MR.

For both BMI and smoking initiation genetic scores there was evidence of assortative mating (Table S9) and associations between these genetic scores and the other health behaviours and household income (Table S10). Additional sensitivity analyses which are more robust to pleiotropy were consistent and there was no evidence for bias from directional horizontal pleiotropy (Table S11). Sensitivity analyses in women including non-planners with a median time to conception suggested an association between higher alcohol consumption and decreased time to conception (−2.653 months, 95% CI: -4.193, -1.112) and between higher BMI and increased time to conception (0.969 months, 95% CI: 0.811, 1.129), that were not observed when restricting to planners (see Supplementary Note 2 for more details).

### Stage 3. Summary-level Mendelian randomisation

There was strong evidence for an effect of smoking initiation on younger age at first birth in women (−0.661, 95% CI: -0.757, -0.566), a greater number of children in both men and women (0.280, 95% CI: 0.205, 0.355) and fewer miscarriages in women (−0.123, 95% CI: -0.182, -0.064) (see Table 2). All pleiotropy robust sensitivity analyses showed consistently strong evidence with the same direction of effect. There was evidence of significant heterogeneity (Table S16) but the MR Egger intercept suggested that these results were not biased by directional pleiotropy (Table S17). Steiger filtering indicated that the majority of SNPs explained more variance in smoking initiation than the outcomes (Table S18), suggesting reverse causation is unlikely.

**Table 2.**
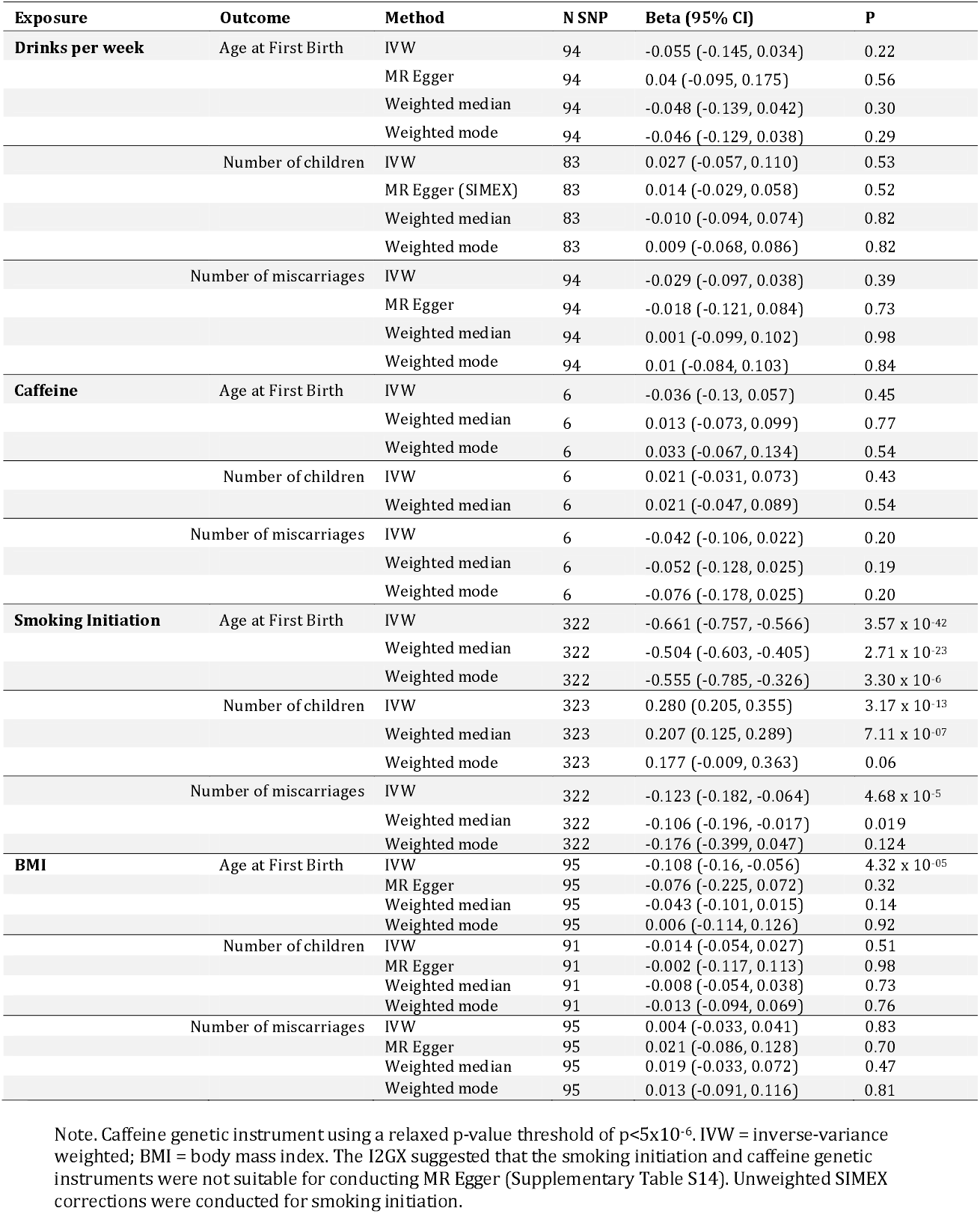
Summary level Mendelian randomisation results for health behaviours on age at first birth, number of children and number of miscarriages.

There was also some evidence for an effect of higher BMI on younger age at first birth in women (−0.108, 95% CI: -0.16, -0.056). All MR sensitivity analyses showed a consistent direction of effect apart from the weighted mode which could indicate possible pleiotropy (Table 2). There was strong evidence of heterogeneity (Table S16), but the MR Egger intercept did not suggest this was due to bias from horizontal pleiotropy (Table S17) and there was no evidence of reverse causation from Steiger filtering (Table S18). There was no evidence for an effect of alcohol consumption or caffeine consumption on reproductive behaviours. All genetic instruments had F-statistic > 10 apart from that for smoking initiation (Table S19).

We conducted multivariable MR analysis (48) to estimate the direct effects of smoking initiation and BMI on age at first birth in women after accounting for education and ADHD liability (see Supplementary Materials for details and Table S20). Effect estimates for smoking initiation attenuated after adjustment but there was still evidence for an effect (after adjusting for ADHD: - 0.435, 95% CI: -0.591, -0.279; after adjusting for education: -0.403, 95% CI:-0.527, -0.279). Effect estimates for BMI were also attenuated, resulting in weak evidence for an effect (ADHD: -0.513, 95% CI: -0.106, 0.003; education: -0.056, 95% CI: -0.113, 0.0008).

## Discussion

This study explored the role of multiple health behaviours on fertility outcomes in men and women using the MoBa cohort. We extended previous research by including men as well as women, and strengthening causal inference using MR. We found evidence from multivariable regression for an association between higher BMI and smoking (and to a lesser extent caffeine) prior to pregnancy on reduced fertility outcomes. Our MR results were not in line with the observational associations, finding only support for an effect of higher BMI and smoking initiation on a younger age at first birth, with evidence of possible horizontal pleiotropy from ADHD liability and education.

Frequency of alcohol consumption was associated with being less likely to have infertility treatment in multivariable regression analyses for both women and men. In contrast, previous meta-analyses have shown a dose-response relationship between alcohol consumption and reduced likelihood of conception in women (5) and reduced semen quality in men (14). However, it is important to note that our analyses were conducted in a pregnancy cohort, so do not capture those who failed to conceive, introducing possible selection bias. Furthermore, levels of alcohol consumption were low in our sample, and are possibly below the threshold which affects fertility (49). To capture more harmful drinking behaviours, we used a measure of binge drinking, which showed weaker associations, supporting our interpretation. It might be feasible that low levels of alcohol consumption are not detrimental for fertility, but it seems unlikely that alcohol could improve fertility, as observed here. Highly confounded multivariable regression associations of alcohol consumption are a common phenomenon, with low levels of consumption often being associated with positive outcomes due to confounding by socio-economic position, or due to never drinkers being a selected group (50). Bias from confounding is supported by our MR analyses, finding no evidence for effects of alcohol consumption on any fertility outcomes. Alternatively, our observational results might be affected by reporting bias, as underreporting of alcohol consumption might be more pronounced amongst those struggling to conceive. Unfortunately, we were unable to follow up binge drinking associations with MR, as the genetic instrument did not predict binge drinking in the MoBa cohort. This is not altogether surprising, as MoBa represents a very different sample to that of the UK Biobank, where the binge drinking GWAS was conducted: 1) the UK Biobank were aged between 40 and 69 years at recruitment (51) compared with an average age of 31 years in MoBa mothers and 33 years in MoBa fathers, 2) response rate was higher for the MoBa cohort (41%) compared to the UK Biobank (5%) (22,52), 3) drinking behaviours differ greatly between the UK and Norway (53), with only 6.31% of MoBa mums reporting binge drinking at least weekly in the 3 months prior to pregnancy, and 4) MoBa is a pregnancy cohort rather than a population cohort, capturing only fertile couples.

There was no evidence for an association between caffeine consumption and miscarriage risk, in contrast to what has been reported in several meta-analyses (54). This could be because our study explored reported caffeine consumption levels prior to pregnancy rather than during pregnancy, which has been the primary focus of most previous meta-analyses (54). Alternatively, it could be due to social patterning of caffeine consumption in Norway, with higher consumption associated with lower levels of education (24) and consequently a younger age at first birth. Older age is a strong predictor of miscarriage risk (55), so education could be masking the association. MR results (which are more robust to bias from confounding) did not find evidence for a causal effect, so it could be possible that previous associations were due to confounding from other lifestyle factors (56). However, it is important to note that the caffeine genetic instrument was the weakest, explaining only 0.1-0.2% of the variance. Due to this weak instrument bias, causal effects cannot be ruled out (57) and replication is warranted.

We found evidence for an association between smoking heaviness and increased time to conception in both women and men. This is supported by a previous meta-analysis finding increased odds of infertility in smokers compared to non-smokers (6). Smoking is hypothesised to negatively affect sperm production and quality in males (58) and to affect the follicles and hormone levels in females (49). However, the effect sizes observed in the current study were quite small and were not supported by the MR results.

Finally, we saw evidence from multivariable regressions for associations between higher BMI and reduced fertility, including taking longer to conceive and an increased risk of miscarriage. These associations are thought to be due to obesity leading to hormone imbalances and ovulatory dysfunction (similar to very low body weights) (59). However, we did not see evidence that this effect was causal in the MR analyses. This could be due to confounding, or due to the MR analysis being unable to detect non-linear effects. Another MR analysis of BMI on subfertility in the MoBa cohort did find evidence for non-linear effects (60). Intervention evidence is mixed, with a meta-analysis including randomised control trials finding evidence that interventions for weight loss increased chance of pregnancy but did not affect risk of miscarriage (61).

A review of studies into health behaviours and fertility concluded that the evidence was robust for smoking and higher weight reducing fertility (49). However, in the current study, our MR results did not find evidence to support this effect. In both the individual-level and summary-level MR we saw consistent evidence for an effect of smoking initiation and higher BMI on having a younger age at first birth. There is also the possibility that these results are due to selection bias: both the GWAS for age at first birth and being a participant in the MoBa sample are conditioned on having had at least one child. If BMI and smoking are associated with reduced fertility, perhaps only those who had children younger were able to conceive and consequently be in the sample. However, if this were the case, we might expect to also see effects of smoking and BMI on other indicators of fertility. An alternative explanation could be that our estimates are biased by horizontal pleiotropy. We used a range of sensitivity analyses to explore potential assumption violation by horizontal pleiotropy. Methods which are agnostic to the specific sources of pleiotropy, (for example, MR-Egger), suggested that the results were not importantly biased by unbalanced horizontal pleiotropy. However, our exploratory multivariable MR analyses did show strong attenuation of the effects of BMI and smoking initiation on age at first birth after accounting for ADHD liability (proxy for ADHD traits of inattentive and hyperactive-impulsive behaviour) and educational attainment (known risk factors for age at first birth). This is likely because ADHD traits affect smoking behaviour and some of the smoking genetic variants relate to smoking via their relationship to ADHD traits. Previous studies have shown a strong association between the smoking initiation instrument and risk-taking behaviours including number of sexual partners (62), which could increase the likelihood of having children younger. Previous Mendelian randomisation studies have also found evidence for bi-directional causal effects between smoking and education (63,64), smoking and ADHD (65), BMI and education (66,67) and BMI and ADHD (68). Bi-directional effects between the exposure and the non-exposure traits can make it difficult to disentangle horizontal from vertical pleiotropy (69). However, several of these previous studies did find evidence of horizontal pleiotropy, especially for the smoking initiation instrument (64,65,68). Therefore, we conclude that horizontal pleiotropy is the most plausible pathway. If there is indeed horizontal pleiotropy from ADHD liability and education, then direct effects from MVMR accounting for these traits will be closer to the true causal effect. Finally, there was also strong evidence for assortative mating for both BMI and smoking initiation instruments which can bias MR results, even for methods which are robust to horizontal pleiotropy (70).

### Strengths and Limitations

The current study has several strengths. The majority of epidemiological research to date has focused on women (13), but we also included fathers in our analysis. We observed a similar pattern of results for women and men in our multivariable regression analyses, despite different hypothesised mechanisms. This supports our interpretation of widespread confounding, because pathways through different mechanisms are unlikely to have the same magnitude of effect (71). Second, this was a very large sample of genotyped individuals with detailed measures of fertility. Third, we combined multivariable regression and MR methods which each rely on different assumptions and therefore triangulating across them can strengthen causal inference. Finally, we included a wide range of health behaviours and different indicators of fertility.

This study does have several limitations. First, all MoBa participants were recruited during pregnancy (12-18 weeks gestation). This means that we are unable to capture the full range of fertility in the population. Those who never managed to conceive were not observed, and this could induce selection bias. Second, multivariable regression analyses were cross-sectional and it is therefore difficult to assess temporality for these associations. Specifically, health behaviours were retrospectively reported about behaviours 3 or 6 months prior to the index pregnancy however some couples had been trying to conceive for longer than 6 months. Furthermore, variables from the Medical Birth Registry of Norway (age at first birth and total number of children) are across all births, and therefore health behaviours may have differed compared to before the index pregnancy. Relatedly, there was also a difference between those in the sample who were planning to conceive compared with those who were not. We hypothesise that planners are more likely to be cautious about their health behaviours, especially if they have been having trouble conceiving and have been advised to quit smoking, stop drinking and lose weight. This could lead to reverse or weakened patterns of association in the multivariable regression analyses. However, MR would be robust to this type of bias, given that genetic propensities to health behaviours are fixed at birth. This might explain the different pattern of results between the multivariable regression and MR analyses. Third, for this paper we have assumed that partners in MoBa were men. In our genetic analyses this is the case, because individuals who were not chromosomally XY were removed from the father analysis. However, in the observational analysis, some partners might have been female partners of the mother, and these individuals could not be identified. Finally, this study conducted triangulation within a single sample, and an approach for stronger causal inference would be to triangulate across different samples in future studies.

## Conclusions

For accurate fertility guidance it is extremely important to establish causality. The majority of associations between health behaviours and fertility observed here, were not replicated using MR methods. This could suggest that previous observational associations were due to confounding or other sources of bias, and consequently improving these health behaviours may not increase the likelihood of conception. We found evidence of potential horizontal pleiotropy, as our genetic instruments for smoking initiation and BMI were also capturing educational attainment and ADHD liability. Therefore, triangulation across a broader range of methods, including those not susceptible to pleiotropy, are required to establish causality.

## Data Availability

The consent and ethical approvals for MoBa does not allow storage of data in repositories or journals, but it is possible to apply for access to summary statistics datasets for replication or reproduction of studies by sending an application to. Data access requires approval from The Regional Committee for Medical and Health Research Ethics in Norway and an agreement with MoBa.

## Funding and Acknowledgements

This work was funded by a Gro Harlem Bruntland Scholarship from the Centre of Fertility and Health NIPH to Dr Wootton. Dr Wootton is currently funded by a postdoctoral fellowship from the South-Eastern Regional Health Authority (2020024). AH was supported by a career grant from the South-Eastern Regional Health Authority (2020022), and by the Research Council of Norway (300668, 288083, 274611). TRK was supported by the Research Council of Norway (274611). PM and SEH were partly funded by the Research Council of Norway (262700 and 320656). GDS and MRM is part of the Medical Research Council Integrative Epidemiology Unit (MRC IEU) at the University of Bristol (MM_UU_00011/1, MM_UU_00011/7). JLT was supported by a Veni grant from the Netherlands Organization for Scientific Research (NWO: grant number 016.Veni.195.016) and by the Foundation Volksbond Rotterdam. RBL is supported by the National Institute of Mental Health grant (R01 MH101269).

This study includes data from the Norwegian Mother, Father and Child Cohort Study (MoBa) conducted by the Norwegian Institute of Public Health. The Norwegian Mother, Father and Child Cohort Study is supported by the Norwegian Ministry of Health and Care Services and the Ministry of Education and Research. We are grateful to all the participating families in Norway who take part in this on-going cohort study. We thank the Norwegian Institute of Public Health (NIPH) for generating high-quality genomic data. This research is part of the HARVEST collaboration, supported by the Research Council of Norway (#229624). For providing genotype data we thank deCODE Genetics, and the NORMENT Centre for providing genotype data, funded by the Research Council of Norway (#223273), South East Norway Health Authority and KG Jebsen Stiftelsen. We further thank the Center for Diabetes Research, the University of Bergen for providing genotype data and performing quality control and imputation of the data funded by the ERC AdG project SELECTionPREDISPOSED, Stiftelsen Kristian Gerhard Jebsen, Trond Mohn Foundation, the Research Council of Norway, the Novo Nordisk Foundation, the University of Bergen, and the Western Norway health Authorities (Helse Vest). This research was also supported by the Research Council or Norway through its Centres of Excellence funding scheme (project No 262700). MCM has funding from the European Research Council (ERC) under the European Union’s Horizon 2020 research and innovation program (grant agreement No 947684).

## Ethical statement

The establishment and data collection in MoBa was previously based on a license from the Norwegian Data protection agency and approval from The Regional Committee for Medical Research Ethics, and it is now based on regulations related to the Norwegian Health Registry Act. The current study was approved by The Regional Committees for Medical and Health Research Ethics (2016/217). All GWAS summary statistics are publicly available.

## References

1. Kamel RM. Management of the infertile couple: an evidence-based protocol. Reprod Biol Endocrinol. 2010 Mar 6;8:21.

2. Norwegian Directorate of Health. Gode levevaner før og i svangerskapet [Internet]. 2018. Available from: https://www.helsedirektoratet.no/brosjyrer/gode-levevaner-for-og-i-svangerskapet/Gode%20levevaner%20f%C3%B8r%20og%20i%20svangerskapet%20-%20engelsk.pdf/_/attachment/inline/73fa64c1-16f3-4052-851c-6a3b2e13ef12:33aa515ffb4a714584b93e4fad70568ec85f4fd0/Gode%20levevaner%20f%C3%B8r%20og%20i%20svangerskapet%20-%20engelsk.pdf

3. Boivin J, Takefman J, Braverman A. The fertility quality of life (FertiQoL) tool: development and general psychometric properties. Hum Reprod Oxf Engl. 2011 Aug;26(8):2084–91.

4. Anderson K, Nisenblat V, Norman R. Lifestyle factors in people seeking infertility treatment–a review. Aust N Z J Obstet Gynaecol. 2010;50(1):8–20.

5. Fan D, Liu L, Xia Q, Wang W, Wu S, Tian G, et al. Female alcohol consumption and fecundability: a systematic review and dose-response meta-analysis. Sci Rep. 2017;7(1):13815.

6. Augood C, Duckitt K, Templeton AA. Smoking and female infertility: a systematic review and meta-analysis. Hum Reprod. 1998 Jun 1;13(6):1532–9.

7. Munafo M, Murphy M, Whiteman D, Hey K. Does cigarette smoking increase time to conception? J Biosoc Sci. 2002;34(1):65–73.

8. Cooper AR, Moley KH. Maternal tobacco use and its preimplantation effects on fertility: more reasons to stop smoking. In: Seminars in reproductive medicine. \copyright Thieme Medical Publishers; 2008. p. 204–12.

9. Wesselink AK, Hatch EE, Rothman KJ, Mikkelsen EM, Aschengrau A, Wise LA. Prospective study of cigarette smoking and fecundability. Hum Reprod Oxf Engl. 2019 Mar 1;34(3):558–67.

10. Cavalcante MB, Sarno M, Peixoto AB, Araujo Júnior E, Barini R. Obesity and recurrent miscarriage: A systematic review and meta-analysis. J Obstet Gynaecol Res. 2019;45(1):30–8.

11. Li J, Zhao H, Song J-M, Zhang J, Tang Y-L, Xin C-M. A meta-analysis of risk of pregnancy loss and caffeine and coffee consumption during pregnancy. Int J Gynecol Obstet. 2015 Aug 1;130(2):116–22.

12. Easey K, Sharp GC. The impact of paternal alcohol, tobacco, caffeine use and physical activity on offspring mental health: A systematic review and meta-analysis. medRxiv. 2021;

13. Sharp GC, Lawlor DA, Richardson SS. It’s the mother!: How assumptions about the causal primacy of maternal effects influence research on the developmental origins of health and disease. Soc Sci Med. 2018;213:20–7.

14. Li Y, Lin H, Li Y, Cao J. Association between socio-psycho-behavioral factors and male semen quality: systematic review and meta-analyses. Fertil Steril. 2011;95(1):116–23.

15. Campbell JM, Lane M, Owens JA, Bakos HW. Paternal obesity negatively affects male fertility and assisted reproduction outcomes: a systematic review and meta-analysis. Reprod Biomed Online. 2015 Nov 1;31(5):593–604.

16. Nguyen RH, Wilcox AJ, Skj\aerven R, Baird DD. Men’s body mass index and infertility. Hum Reprod. 2007;22(9):2488–93.

17. Peck JD, Leviton A, Cowan LD. A review of the epidemiologic evidence concerning the reproductive health effects of caffeine consumption: A 2000–2009 update. Food Chem Toxicol. 2010 Oct 1;48(10):2549–76.

18. Groenman AP, Janssen TWP, Oosterlaan J. Childhood Psychiatric Disorders as Risk Factor for Subsequent Substance Abuse: A Meta-Analysis. J Am Acad Child Adolesc Psychiatry. 2017 Jul 1;56(7):556–69.

19. Kong A, Frigge ML, Thorleifsson G, Stefansson H, Young AI, Zink F, et al. Selection against variants in the genome associated with educational attainment. Proc Natl Acad Sci. 2017;114(5):E727–32.

20. Hvolgaard Mikkelsen S, Olsen J, Bech BH, Obel C. Parental age and attention-deficit/hyperactivity disorder (ADHD). Int J Epidemiol. 2017 Apr 1;46(2):409–20.

21. Davey Smith G, Ebrahim S. “Mendelian randomization”: can genetic epidemiology contribute to understanding environmental determinants of disease? Int J Epidemiol. 2003 Feb;32(1):1–22.

22. Magnus P, Irgens LM, Haug K, Nystad W, Skj\a erven R, Stoltenberg C. Cohort profile: the Norwegian mother and child cohort study (MoBa). Int J Epidemiol. 2006;35(5):1146–50.

23. Magnus P, Birke C, Vejrup K, Haugan A, Alsaker E, Daltveit AK, et al. Cohort profile update: the Norwegian mother and child cohort study (MoBa). Int J Epidemiol. 2016;45(2):382–8.

24. Papadopoulou E, Botton J, Brants\a eter A-L, Haugen M, Alexander J, Meltzer HM, et al. Maternal caffeine intake during pregnancy and childhood growth and overweight: results from a large Norwegian prospective observational cohort study. BMJ Open. 2018;8(3):e018895.

25. Treur JL, Taylor AE, Ware JJ, McMahon G, Hottenga J-J, Baselmans BM, et al. Associations between smoking and caffeine consumption in two European cohorts. Addiction. 2016;111(6):1059–68.

26. Paltiel L, Anita H, Skjerden T, Harbak K, Bækken S, Kristin SN, et al. The biobank of the Norwegian Mother and Child Cohort Study – present status. Nor Epidemiol [Internet]. 2014 Dec 22 [cited 2021 Jun 9];24(1–2). Available from: https://www.ntnu.no/ojs/index.php/norepid/article/view/1755

27. Helgeland Ø, Vaudel M, Juliusson PB, Lingaas Holmen O, Juodakis J, Bacelis J, et al. Genome-wide association study reveals dynamic role of genetic variation in infant and early childhood growth. Nat Commun. 2019 Oct 1;10(1):4448.

28. Euesden J, Lewis CM, O’Reilly PF. PRSice: Polygenic Risk Score software. Bioinforma Oxf Engl. 2015 May 1;31(9):1466–8.

29. Liu M, Jiang Y, Wedow R, Li Y, Brazel DM, Chen F, et al. Association studies of up to 1.2 million individuals yield new insights into the genetic etiology of tobacco and alcohol use. Nat Genet. 2019;51:237–44.

30. Cornelis MC, Byrne EM, Esko T, Nalls MA, Ganna A, Paynter N, et al. Genome-wide meta-analysis identifies six novel loci associated with habitual coffee consumption. Mol Psychiatry. 2015 May;20(5):647–56.

31. Jensen KP, DeVito EE, Herman AI, Valentine GW, Gelernter J, Sofuoglu M. A CHRNA5 Smoking Risk Variant Decreases the Aversive Effects of Nicotine in Humans. Neuropsychopharmacology. 2015 Nov;40(12):2813–21.

32. Yengo L, Sidorenko J, Kemper KE, Zheng Z, Wood AR, Weedon MN, et al. Meta-analysis of genome-wide association studies for height and body mass index in ∼700,000 individuals of European ancestry. bioRxiv. 2018 Mar 2;274654.

33. R. Core Team. R: A language and environment for statistical computing. R Foundation for Statistical Computing, Vienna, Austria. 2013. ISBN 3-900051-07-0; 2014.

34. Kessler RC, Adler L, Ames M, Demler O, Faraone S, Hiripi E, et al. The World Health Organization Adult ADHD Self-Report Scale (ASRS): a short screening scale for use in the general population. Psychol Med. 2005 Feb;35(2):245–56.

35. Lousdal ML. An introduction to instrumental variable assumptions, validation and estimation. Emerg Themes Epidemiol. 2018 Jan 22;15:1.

36. Bowden J, Davey Smith G, Burgess S. Mendelian randomization with invalid instruments: effect estimation and bias detection through Egger regression. Int J Epidemiol. 2015;44(2):512–25.

37. Bowden J, Davey Smith G, Haycock PC, Burgess S. Consistent estimation in Mendelian randomization with some invalid instruments using a weighted median estimator. Genet Epidemiol. 2016;40(4):304–14.

38. Hartwig FP, Smith GD, Bowden J. Robust inference in two-sample Mendelian randomisation via the zero modal pleiotropy assumption. bioRxiv. 2017;126102.

39. Purcell S, Neale B, Todd-Brown K, Thomas L, Ferreira MA, Bender D, et al. PLINK: a tool set for whole-genome association and population-based linkage analyses. Am J Hum Genet. 2007;81(3):559–75.

40. Lawlor DA. Commentary: Two-sample Mendelian randomization: opportunities and challenges. Int J Epidemiol. 2016;45(3):908.

41. Sudlow C, Gallacher J, Allen N, Beral V, Burton P, Danesh J, et al. UK biobank: an open access resource for identifying the causes of a wide range of complex diseases of middle and old age. PLoS Med. 2015;12(3):e1001779.

42. Elsworth B, Lyon M, Alexander T, Liu Y, Matthews P, Hallett J, et al. The MRC IEU OpenGWAS data infrastructure. bioRxiv. 2020 Aug 10;2020.08.10.244293.

43. Lawn RB, Sallis HM, Taylor AE, Wootton RE, Smith GD, Davies NM, et al. Schizophrenia risk and reproductive success: A Mendelian randomization study. R Soc Open Sci. 2018 Jun 28;357673.

44. Locke AE, Kahali B, Berndt SI, Justice AE, Pers TH, Day FR, et al. Genetic studies of body mass index yield new insights for obesity biology. Nature. 2015;518(7538):197–206.

45. Burgess S, Davies NM, Thompson SG. Bias due to participant overlap in two-sample Mendelian randomization. Genet Epidemiol. 2016 Nov;40(7):597–608.

46. Bowden J, Del Greco M F, Minelli C, Davey Smith G, Sheehan NA, Thompson JR. Assessing the suitability of summary data for two-sample Mendelian randomization analyses using MR-Egger regression: the role of the I 2 statistic. Int J Epidemiol. 2016;45(6):1961–74.

47. Hemani G, Tilling K, Davey Smith G. Orienting the causal relationship between imprecisely measured traits using GWAS summary data. Li J, editor. PLOS Genet. 2017 Nov 17;13(11):e1007081.

48. Sanderson E, Davey Smith G, Windmeijer F, Bowden J. An examination of multivariable Mendelian randomization in the single-sample and two-sample summary data settings. Int J Epidemiol. 2019;48(3):713–27.

49. Homan GF, Davies M, Norman R. The impact of lifestyle factors on reproductive performance in the general population and those undergoing infertility treatment: a review. Hum Reprod Update. 2007 May 1;13(3):209–23.

50. Connor J. The Life and Times of the J-Shaped Curve. Alcohol Alcohol. 2006 Nov 1;41(6):583–4.

51. Bycroft C, Freeman C, Petkova D, Band G, Elliott LT, Sharp K, et al. The UK Biobank resource with deep phenotyping and genomic data. Nature. 2018;562(7726):203–9.

52. Munafò MR, Tilling K, Taylor AE, Evans DM, Davey Smith G. Collider scope: when selection bias can substantially influence observed associations. Int J Epidemiol. 2018 Feb 1;47(1):226–35.

53. World Health Organization. Global status report on alcohol and health 2018 [Internet]. Geneva; 2018 [cited 2019 Jan 4]. Available from: http://apps.who.int/iris/bitstream/handle/10665/274603/9789241565639-eng.pdf?ua=1.

54. James JE. Maternal caffeine consumption and pregnancy outcomes: a narrative review with implications for advice to mothers and mothers-to-be. BMJ Evid-Based Med. 2021 Jun 1;26(3):114–5.

55. Magnus MC, Wilcox AJ, Morken N-H, Weinberg CR, Håberg SE. Role of maternal age and pregnancy history in risk of miscarriage: prospective register based study. BMJ. 2019 Mar 20;364: l869.

56. Chen L, Bell EM, Browne ML, Druschel CM, Romitti PA, National Birth Defects Prevention Study. Exploring maternal patterns of dietary caffeine consumption before conception and during pregnancy. Matern Child Health J. 2014 Dec;18(10):2446–55.

57. Cornelis MC, Munafo MR. Mendelian Randomization Studies of Coffee and Caffeine Consumption. Nutrients. 2018 Oct;10(10):1343.

58. Künzle R, Mueller MD, Hänggi W, Birkhäuser MH, Drescher H, Bersinger NA. Semen quality of male smokers and nonsmokers in infertile couples. Fertil Steril. 2003 Feb 1;79(2):287–91.

59. Silvestris E, de Pergola G, Rosania R, Loverro G. Obesity as disruptor of the female fertility. Reprod Biol Endocrinol. 2018 Mar 9;16(1):22.

60. Hernáez Á, Rogne T, Skåra KH, Håberg SE, Page CM, Fraser A, et al. Body mass index and subfertility: multivariable regression and Mendelian randomization analyses in the Norwegian Mother, Father and Child Cohort Study. Hum Reprod. 2021 Dec 1;36(12):3141–51.

61. Best D, Avenell A, Bhattacharya S. How effective are weight-loss interventions for improving fertility in women and men who are overweight or obese? A systematic review and meta-analysis of the evidence. Hum Reprod Update. 2017 Nov 1;23(6):681–705.

62. Khouja JN, Wootton RE, Taylor AE, Smith GD, Munafò MR. Association of genetic liability to smoking initiation with e-cigarette use in young adults: A cohort study. PLOS Med. 2021 Mar 18;18(3):e1003555.

63. Gage SH, Bowden J, Davey Smith G, Munafò MR. Investigating causality in associations between education and smoking: a two-sample Mendelian randomization study. Int J Epidemiol. 2018 Aug 1;47(4):1131–40.

64. Gage SH, Sallis HM, Lassi G, Wootton RE, Mokrysz C, Davey Smith G, et al. Does smoking cause lower educational attainment and general cognitive ability? Triangulation of causal evidence using multiple study designs. Psychol Med. 2020 Oct 7;1–9.

65. Treur JL, Demontis D, Smith GD, Sallis H, Richardson TG, Wiers RW, et al. Investigating causality between liability to ADHD and substance use, and liability to substance use and ADHD risk, using Mendelian randomization. Addict Biol. 2021 Jan;26(1):e12849.

66. Böckerman P, Viinikainen J, Pulkki-Råback L, Hakulinen C, Pitkänen N, Lehtimäki T, et al. Does higher education protect against obesity? Evidence using Mendelian randomization. Prev Med. 2017 Aug;101:195–8.

67. Hughes A, Wade KH, Dickson M, Rice F, Davies A, Davies NM, et al. Common health conditions in childhood and adolescence, school absence, and educational attainment: Mendelian randomization study. Npj Sci Learn. 2021 Jan 4;6(1):1–9.

68. Liu C-Y, Schoeler T, Davies NM, Peyre H, Lim K-X, Barker ED, et al. Are there causal relationships between attention-deficit/hyperactivity disorder and body mass index? Evidence from multiple genetically informed designs. Int J Epidemiol. 2021 May 17;50(2):496–509.

69. Yang Q, Sanderson E, Tilling K, Borges MC, Lawlor DA. Exploring and mitigating potential bias when genetic instrumental variables are associated with multiple non-exposure traits in Mendelian randomization [Internet]. medRxiv; 2019 [cited 2022 Feb 17]. p. 19009605. Available from: https://www.medrxiv.org/content/10.1101/19009605v1

70. Hartwig FP, Davies NM, Davey Smith G. Bias in Mendelian randomization due to assortative mating. Genet Epidemiol. 2018 Oct;42(7):608–20.

71. Smith GD. Assessing Intrauterine Influences on Offspring Health Outcomes: Can Epidemiological Studies Yield Robust Findings? Basic Clin Pharmacol Toxicol. 2008;102(2):245–56.

