## Supplementary Materials for "Health behaviours prior to pregnancy and fertility outcomes: Triangulation of evidence in the Norwegian Mother, Father and Child Cohort Study (MoBa)"

**Contents.**

| Supplementary Figures  S1. An overview of participant inclusion and attrition.  S2. The influence of rs16969968 genotype on fertility outcomes  Supplementary Notes | Page 2  Page 3 |
| --- | --- |
| Note 1. Genotyping and Quality control procedures | Page 4 |
| Note 2. Comparison between planning and non-planning couples | Page 7 |
| Note 3. Sensitivity analysis dichotomising time to conception | Page 8 |
| Note 4. Health behaviours and frequency of sexual intercourse | Page 10 |
| Note 5. Single-SNP analysis of rs16969968 genotype stratified by smoking status | Page 11 |
| Supplementary Tables |  |
| S1. Testing instrument strength of each of the polygenic risk scores | Page 12 |
| S2. Associations between fertility outcomes  S3. Observational associations between health behaviours in the mothers | Page 13  Page 14 |
| S4. Observational associations between health behaviours in the fathers | Page 15 |
| S5. Observational associations between health behaviours in the mothers genotyped sample only | Page 16 |
| S6. Observational associations between health behaviours in the fathers genotyped sample only | Page 18 |
| S7. Individual-level Mendelian randomisation analysis in the mothers | Page 19 |
| S8. Individual-level Mendelian randomisation analysis in the fathers | Page 20 |
| S9. Evidence for assortative mating | Page 21 |
| S10. Evidence for reintroduced confounding | Page 22 |
| S11. Summary level MR sensitivity tests conducted in the MoBa sample | Page 23 |
| S12. Summary of mother characteristics comparing planners and non-planners | Page 24 |
| S13. Including non-planners in time to conception observational analysis | Page 25 |
| S14. Including non-planners in time to conception MR | Page 26 |
| S15. Frequency of self-reported sexual intercourse | Page 27 |
| S16. Evidence for heterogeneity: Cochran’s Q statistics | Page 28 |
| S17. The MR Egger intercept test: Evidence for bias from horizontal pleiotropy | Page 29 |
| S18. Steiger filtering test for possible reverse causation | Page 30 |
| S19. Test of instrument strength and the suitability of the instrument for MR Egger | Page 31 |
| S20. Exploratory multivariable Mendelian randomisation | Page 32 |
| References | Page 33 |

**Figure S1. An overview of participant inclusion and attrition.**

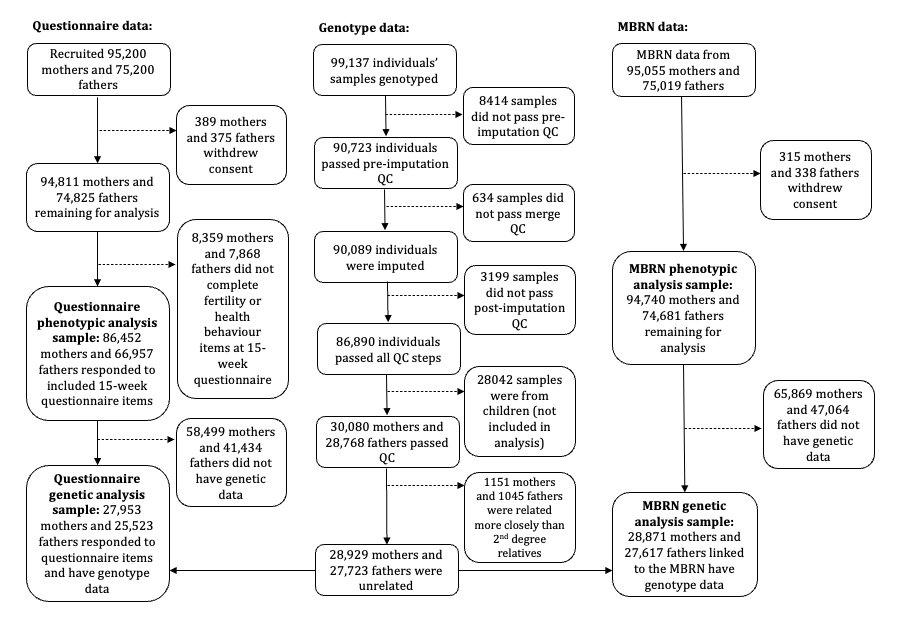

**Note.** QC = quality control, MBRN = medical birth registry of Norway. Analysis samples represent the maximum number of individuals available for analysis. For specific N for each variable, see Table 1 in the main manuscript. Variables from the MBRN were parental age, parent’s age at first birth and total number of children. All other variables were from questionnaire data.

**Figure S2. The influence of rs16969968 genotype (A allele) on fertility outcomes stratified by smoking status prior to pregnancy**

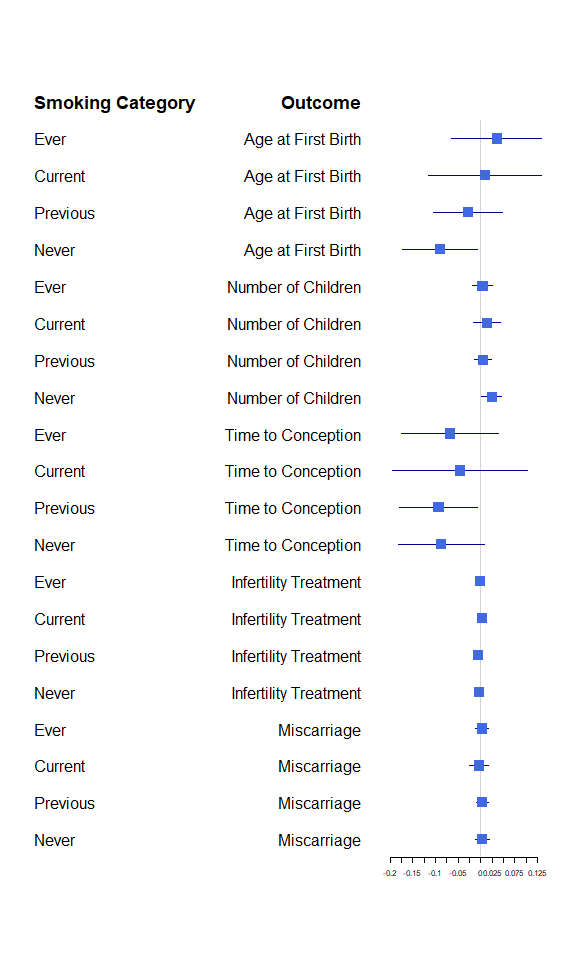

**Supplementary Note 1. Genotyping and Quality control procedures**

Genotype Data

For the current study we used MoBa Genetics genotype data release 1.0, based on genotype data from four research projects. This release has genotype data available for 98,110 individuals (mostly family trios). 33,199 individuals from the NORMENT project were genotyped at deCODE genetics, Reykjavik Iceland (Illumina HumanOmniExpress-24v1.0, Illumina InfiniumOmniExpress-24v1.2, & Illumina Global Screening Array MD v.1.0 + 50k custom OmniExpress overlap content array), 26,990 were genotyped in the ROTTERDAM project at ERASMUS MC, Rotterdam, Netherlands (Illumina Global Screening Array MD v.1.0 array), and 5,410 from the ADHD project (TED) were genotyped at deCODE genetics (Illumina InfiniumOmniExpress-24v1.2), and 32,538 were sampled in the HARVEST sample at Genomics Core Facility, Trondheim, Norway (llumina HumanCoreExome12v1.1 & Illumina HumanCoreExome24v1.0). Pre-imputation QC, phasing and imputation are described elsewhere (Helgeland et al., 2021).

Post-imputation data quality control

Individuals with sex-mismatch (derived by comparing genetic sex and reported sex) or individuals with sex-chromosome aneuploidy or those that were not linkable with the phenotypic data were excluded from the analysis (n=508). We checked the dataset for Mendelian errors using PLINK’s --mendel command with thresholds of 1% and 5% for the trio and variant error rate respectively. This excluded 129 individuals and 1293 variants. On average these variants were less precisely imputed, mean INFO=0.88 versus mean INFO=0.97 for the rest of the SNPs.

3,061 individuals were genotyped twice, and 52 individuals were genotyped 3 times. We checked the concordance of SNPs within pairs of duplicated samples, and 2 samples, that were indicated as duplicates by the MoBa data, had very low concordance (pi_hat<0.03), and were excluded. The remaining individuals had high concordance (pi_hat>0.8). Note there were some 2,474 pairs of samples that had concordance between 0.74 and 0.98. These individuals are almost certainly the same individuals and are unlikely to represent sample contamination (this would have been picked up at the genotype calling stage). Therefore, this heterogeneity is likely due to the fact that samples for the same individuals are combined from different chips which have not been imputed to the same standard. We excluded 140,767 SNPs which were discordant for more than 5% of duplicated samples. One individual from each pair of duplicates was then dropped at random with a seed.

Ancestry

We restricted the sample to individuals of ‘european’ ancestry using the first 2 principal components of the MoBa data. The principal components were calculated by merging the MoBa data with the 1000g reference panel and projecting the 1000g PCs onto the MoBa data. We then compared the PC values in the MoBa samples versus each of the populations included in the reference panel. We excluded samples if they had values of PC1 and PC2 that were within the range of the non-European samples in the 1000g reference panel, leading to the exclusion of 668 non-European samples.

Degree of relatedness

Estimated kinship coefficients using the KING with age to the nearest year included as a covariate identified 86,175 pairs of known related individuals from the pedigree and a further 10,769 unknown related individuals.

Parental relationships were updated on the basis of KING results. Before running KING, we restricted it to an independent set of high frequency SNPs (MAF>0.10, window=3000kb and LD R2 >0.9). KING estimates the relationships of all the individuals in the datasets and reconstructs families. The output is a list of family and within family IDs. KING updated family IDs for 24,022 individuals, and parental relationships for 21,361 individuals.  Samples were flagged for exclusion if newly assigned relationships appeared to be errors (375 samples). This included where: parents were less than 15 years older than children, both parents in a family were of the same sex, individuals were identified as MZ twins but linked to different pregnancies, and siblings had different parents or an age gap of more than 25 years. We also flagged for exclusion parent-offspring pairs where the mother or father as identified by KING was different to the genotyped individual specified in the pedigree. This category will include both samples where the sampled partner is not the biological father, and potentially sample mix-ups. As our current study analysed mothers and fathers separately, we separated mothers and fathers genotypes and used the KING software (Manichaikul et al., 2010) to identify a subset of mothers and a subset of fathers less similar to one another than 2^nd^ degree relatives. We kept only this subset of unrelated individuals for each analysis. We also checked that this had removed relatedness captured by the other flag lists. The remaining samples contained 28,929 mothers and 27,723 fathers.

Principal components

The first 20 principal components were calculated independently for offspring and parents on a subset of the data limited to variants in HAPMAP3, that were pruned for independence using PLINK. We constructed two sets of principal components, the first used the MoBa data and accounts for structure within the data. The second set of principal components was generated using the 1000 genomes reference panel.

**Supplementary Note 2. Comparison between planners and non-planners**

Methods

Individuals who did not plan their pregnancy cannot provide a time to conception, as this will be unknown. In our primary analyses of time to conception, we have therefore excluded non-planners and included the planners only. Here we explore the difference between the couples who did and didn’t plan their pregnancy. First, we explore the demographic differences between the planning and the non-planning group. Second, we repeated our observational analysis and our individual-level MR analysis on time to conception but with the non-planners included. Non-planners cannot report time to conception, so we gave them the median time to conception from the planning group (2 months).

Results

There are differences in demographics and health behaviours between the planning and the non-planning group (see Supplementary Table S12). The non-planners have more children, have their first child younger, are less likely to be university educated, have lower BMI, smoke more, are more likely to binge drink and drink more caffeine. Therefore, bias will be introduced by selecting the planning group only.

In Supplementary Table S13, we compare the primary observational results with an analysis where non-planners are included. There are disparities between the results. The associations between smoking status and smoking heaviness with increased time to conception were attenuated when including the non-planners. The associations between higher alcohol consumption and binge drinking with reduced time to conception were strengthened when including the non-planners. Results remained relatively consistent for BMI and caffeine consumption.

In Supplementary Table S14, we compare the individual-level MR results, with planners and non-planners included. When including only planners in analysis, there was no evidence for an effect of any of the health behaviours on time to conception using individual-level MR. After the planners were included, there was evidence for genetically predicted higher alcohol consumption on a decreased time to conception (risk difference per SD increase in genetic score: -2.653, 95% CI: -4.193, -1.112) and evidence for genetically predicted higher BMI on increased time to conception (risk difference per SD increase in genetic score: 0.969, 95% CI: 0.811, 1.129).

**Supplementary Note 3. Sensitivity analysis dichotomising time to conception**

Methods

In our primary analysis we used self-reported time to conception as an outcome variable. However, this measure was not linear, with response categories of less than one month, 1-2 months or 3+ months. If it took more than 3 months, then mothers were asked to state the number of months. We combined anyone taking 12 or more months to conceive into one group to reduce skewness and treated as a continuous variable. Given the skew in this variable, we performed a sensitivity analysis where we created two additional dichotomised variables: subfertility and high fertility.

*Subfertility****.*** Subfertility was defined as taking 12 months or longer to conceive. This was based upon mothers self-reported time to conception from the 15-week questionnaire. We have used this mother-reported variable as an outcome in both mothers and fathers as couples were conceiving together.

*High fertility.* A second binary variable represented the other end of the fertility distribution. ‘Highly fertile’ couples were defined as conceiving in less than one month (if they were trying to conceive) or those who conceived despite using contraception (using any contraception methods apart from withdrawal or safe period unless in combination with another form of contraception). This was based upon mothers self-reported time to conception from the 15-week questionnaire. We have used this mother-reported variable as an outcome in both mothers and fathers as couples were conceiving together.

Results

*Descriptives.* 9.9% of mothers and fathers met criteria for subfertility in the full sample and 9.1% in the genotyped sample. 23.1% of mothers and fathers in the full sample met criteria for high fertility compared with 23.8% of the genotyped sample.

*Observational associations.* In mother’s adjusted analyses, greater frequency of alcohol consumption was weakly associated with being more likely to have high fertility (OR = 1.021, 95% CI: 1.004, 1.037) but not with subfertility or with either measure in fathers. Higher BMI in both mothers and fathers was associated with being more likely to have subfertility (mothers OR: 1.048, 95% CI: 1.042, 1.054; fathers OR: 1.037, 95% CI: 1.028, 1.046) and less likely to have high fertility (mothers OR: 0.988, 95% CI: 0.984, 0.992; fathers OR: 0.982, 95%CI: 0.976, 0.988). In mothers nor fathers, higher caffeine consumption and smoking initiation were not associated with neither being highly fertile or odds of subfertility. Higher smoking heaviness was associated with a higher likelihood of subfertility in both women (OR: 1.029, 95% CI: 1.019, 1.040) and men (OR: 1.019, 95% CI: 1.008, 1.030) (Tables S3-S6).

*Individual-level MR.* We found no evidence for association between any of the health behaviours with either high fertility or subfertility after correcting for multiple testing (Tables S7 and S8).

**Supplementary Note 4. Exploring the association between health behaviours and frequency of sexual intercourse**

Methods

As a secondary outcome, we looked at the association between health behaviours and frequency of sexual intercourse. This is not a fertility outcome per se but could be a possible mediator of effects on fertility outcomes (e.g., time to conception, number of children).

*Variable definition.* Mothers self-reported (in the 15-week questionnaire) the frequency they had sexual intercourse in the 4 weeks prior to pregnancy. Response categories were on a 7-point scale from *never* to *every day*. Fathers were assigned the mother’s self-reported frequency if they reported having been in a sexual relationship with the mother for 5 weeks or longer (to ensure they were together for 4 weeks prior to pregnancy). Otherwise, father’s frequency was set to missing.

Results

*Descriptives.* It was most common to have sex 1-2 times a week for both men (36.00%) and women (35.84%). Full response frequencies are given in Table S15.

*Observational associations.* In adjusted analyses, higher frequency of alcohol consumption and binge drinking were both associated with having sex more frequently in women (alcohol frequency: 0.072, 95% CI: 0.065, 0.079; binge drinking: 0.066, 95% CI: 0.057, 0.075) and in men (alcohol frequency: 0.042, 95% CI: 0.034, 0.050; binge drinking: 0.076, 95% CI: 0.061, 0.090). Higher caffeine consumption was associated with increased sex frequency in women (0.018, 95% CI: 0.011, 0.024) but not men. Higher BMI in both men and women was associated with having sex less frequently (women: -0.007, 95% CI: -0.008, -0.005; men: -0.007, 95% CI: -0.010, -0.005). Smoking heaviness was associated with having more frequent sexual intercourse in women (0.008, 95% CI: 0.005, 0.011) and men (0.006, 95% CI: 0.002, 0.009) (Tables S2-S5).

*Individual-level Mendelian randomisation.* Genetic liability for any of the health behaviours was not associated with frequency of sexual intercourse (Tables S7 and S8).

**Supplementary Note 5. Single-SNP analysis of rs16969968 genotype stratified by smoking status**

Methods

We conducted an individual-level MR analysis using a single-nucleotide polymorphism (SNP) for smoking heaviness. We used the genetic variant rs16969968 (A/G) found in the gene cluster *CHRNA5-A3-B4*, a nicotinic receptor subunit gene cluster on chromosome 15 that has been robustly associated with smoking heaviness in smokers (Ducci et al., 2011; Munafò et al., 2012). This SNP is functional and leads to an amino acid change (D398N) in the nicotinic receptor α5 subunit protein (Fowler et al., 2011). The minor allele is associated with nicotine metabolism such that each allele increase in rs16969968 corresponds to an average of one more cigarette smoked per day (Munafò et al., 2012; Thorgeirsson et al., 2008; Tobacco Consortium, 2010). Therefore, if there were to be evidence for an effect in current smokers, this could suggest a causal role of smoking. An additional strength of this approach is being able to compare the effects in current smokers with the effects in never smokers. If there is any evidence for an effect in the never smokers, then this is indicative of horizontal pleiotropy, through pathways other than smoking (Millard et al., 2019).

Genotype at rs16969968 was coded as 0, 1 or 2, counting the number of minor alleles (A). We stratified our sample into ever smokers, previous smokers, current smokers and never smokers, as reported 3-months prior to pregnancy. A linear regression was run of rs16969968 genotype for each smoking status on each continuous fertility outcome and a logistic regression was run for each binary outcome. Analysis was conducted in R, version 4.0.3.

Results

The results of the single SNP analysis are presented below in Supplementary Figure S2. There was no evidence for an effect of smoking heaviness on any of the fertility outcomes, which would be indicated through an association in the current smokers and no association in the never smokers (the negative control). Results are presented as betas for continuous outcomes (per allele increase) and as risk differences for binary outcomes (per allele increase).

**Supplementary Table S1. Testing instrument strength of each of the polygenic risk scores**

| **PRS** | **Exposure** | **N SNP** | **F Statistic (df)** | **R2/Pseudo R2** | **Beta/OR (95% CI)** | **P-value** |
| --- | --- | --- | --- | --- | --- | --- |
| **Mothers** |  |  |  |  |  |  |
| Binge p<5x10-6 | Binge Drinking | 12 | F(1, 26544)=6.88 | 0.0003 | 0.014 (0.004, 0.025) | 0.009 |
| Binge p<5x10-8 | Binge Drinking | 4 | F(1, 26544)=4.56 | 0.0001 | 0.012 (0.001, 0.023) | 0.033 |
| BMI p<5x10-8 | BMI | 902 | F(1, 27270)=1862.54 | 0.064 | 1.078 (1.029, 1.127) | 0.00e+00 |
| Caffeine p<5x10-6 | Caffeine Consumption | 14 | F(1, 24510)=34.12 | 0.001 | 0.045 (0.030, 0.060) | 5.25e-09 |
| Caffeine p<5x10-8 | Caffeine Consumption | 5 | F(1, 24510)=53.67 | 0.002 | 0.057 (0.042, 0.072) | 2.45e-13 |
| CPD p<5x10-8 | Smoking Heaviness | 24 | F(1, 4959)=62.16 | 0.012 | 0.641 (0.482, 0.801) | 3.86e-15 |
| DPW p<5x10-8 | Alcohol Frequency | 49 | F(1, 26817)=21.74 | 0.0008 | 0.031 (0.018, 0.044) | 3.13e-06 |
| SI p<5x10-8 | Smoking Initiation | 187 | - | 0.007 | 1.221 (1.192, 1.251) | 8.59e-60 |
| **Fathers** |  |  |  |  |  |  |
| Binge p<5x10-6 | Binge Drinking | 12 | F(1, 10759)=0.04 | 0.000003 | 0.002 (-0.015, 0.019) | 0.844 |
| Binge p<5x10-8 | Binge Drinking | 4 | F(1, 10759)=1.70 | 0.000 | 0.011 (-0.006, 0.028) | 0.192 |
| BMI p<5x10-8 | BMI | 902 | F(1, 26159)=1640.01 | 0.059 | 0.806 (0.767, 0.846) | 0.00e+00 |
| Caffeine p<5x10-6 | Caffeine Consumption | 14 | F(1, 10324)=25.47 | 0.002 | 0.061 (0.038, 0.085) | 4.56e-07 |
| Caffeine p<5x10-8 | Caffeine Consumption | 5 | F(1, 10324)=17.55 | 0.002 | 0.051 (0.027, 0.074) | 2.82e-05 |
| CPD p<5x10-8 | Smoking Heaviness | 24 | F(1, 4719)=45.67 | 0.010 | 0.598 (0.425, 0.772) | 1.57e-11 |
| DPW p<5x10-8 | Alcohol Frequency | 49 | F(1, 24773)=32.41 | 0.001 | 0.040 (0.026, 0.053) | 1.26e-08 |
| SI p<5x10-8 | Smoking Initiation | 187 |  | 0.005 | 1.180 (1.151, 1.210) | 2.16e-38 |

Note. Variance explained is presented as R^2^ for continuous exposures and as pseudo R^2^ for binary exposures. SI = smoking initiation, DPW = drinks per week, CPD = cigarettes per day, BMI = body mass index, N SNP = the number of SNPs available in the sample to construct the score after clumping.

**Supplementary Table S2. Associations between fertility outcomes in the full sample.**

| **Mothers** | AFB (years) | N Children | TTC (months) | Sex Freq | Miscarriage | Infertility Treatment |
| --- | --- | --- | --- | --- | --- | --- |
| Age at first birth |  | 94,740 | 63,024 | 83,423 | 29,452 | 9,042 |
| Number of children | r=-0.34  p<0.001 |  | 63,024 | 83,423 | 26,792 | 9,042 |
| Time to Conception | r=0.11  p<0.001 | r=-0.08  p<0.001 |  | 62,200 | 19,118 | 4,853 |
| Frequency of sexual intercourse | r=-0.17  p<0.001 | r=0.04  p<0.001 | r=-0.08  p<0.001 |  | 30,897 | 8,871 |
| Miscarriage | Never = 26.56  Ever = 27.55  p<0.001 | Never = 2.68  Ever = 2.75  p<0.001 | Never = 4.18  Ever = 5.57  p<0.001 | Never = 4.28  Ever = 4.34  p<0.001 |  | 55,094 |
| Infertility treatment | Never = 27.05  Ever = 29.78  p<0.001 | Never = 2.55  Ever = 2.36  p<0.001 | Never = 3.81  Ever = 18.03  p<0.001 | Never = 4.41  Ever = 4.08  p<0.001 | $\chi^{2}$= 566 (df=1)  p<0.001 |  |
| **Fathers** | AFB | N Children | TTC | Sex Freq |  |  |
| Age at first birth |  | 74,681 | 53,432 | 67,236 |  |  |
| Number of children | r=-0.36  p<0.001 |  | 53,432 | 67,236 |  |  |
| Time to Conception | r=0.10  p<0.001 | r=-0.06  p<0.001 |  | 52,519 |  |  |
| Frequency of sexual intercourse | r=-0.13  p<0.001 | r=0.02  p<0.001 | r=-0.09  p<0.001 |  |  |  |

Note. We used continuous measures where possible (rather than the categorised measures used in the main analysis). Continuous measures were associated using Pearson’s correlation. Binary variables were associated with continuous measures using independent samples t-test. Binary measures were associated with each other using chi-squared test. Below the diagonal is the Pearson’s correlation coefficient and p-value for correlations, the group means and t-test p-value for t-tests and chi-squared statistic and p-value for chi-squared tests. Above the diagonal is the number of individuals available for each comparison.

**Supplementary Table S3. Observational associations between health behaviours in the mothers (full sample)**

|  |  |  | Unadjusted |  | Adjusted for birthyear and education | |
| --- | --- | --- | --- | --- | --- | --- |
| **Exposure** | **Outcome** | **N** | **Beta (95% CI)** | **P-value** | **Beta (95% CI)** | **P-value** |
| Alcohol Frequency | Age at First Birth | 79505 | 0.898 ( 0.869, 0.926) | 0.00e+00 | 0.492 ( 0.468, 0.516) | 0.00e+00 |
| Alcohol Frequency | Parity | 79505 | -0.094 (-0.100, -0.089) | 1.43e-247 | -0.090 (-0.096, -0.085) | 3.95e-207 |
| Alcohol Frequency | Sex Frequency | 77836 | 0.043 ( 0.036, 0.050) | 2.06e-32 | 0.072 ( 0.065, 0.079) | 9.36e-82 |
| Alcohol Frequency | Time to Conception | 60269 | -0.029 (-0.054, -0.003) | 0.026 | -0.028 (-0.054, -0.001) | 0.041 |
| Binge Drinking | Age at First Birth | 78558 | -0.069 (-0.105, -0.034) | 1.08e-04 | 0.333 ( 0.304, 0.362) | 2.58e-112 |
| Binge Drinking | Parity | 78558 | -0.115 (-0.122, -0.108) | 7.12e-253 | -0.111 (-0.118, -0.104) | 1.67e-220 |
| Binge Drinking | Sex Frequency | 76941 | 0.101 ( 0.092, 0.109) | 1.87e-116 | 0.066 ( 0.057, 0.075) | 2.56e-49 |
| Binge Drinking | Time to Conception | 59677 | -0.028 (-0.059, 0.003) | 0.080 | 0.013 (-0.019, 0.045) | 0.415 |
| Caffeine Consumption | Age at First Birth | 75117 | 0.312 ( 0.285, 0.339) | 4.68e-111 | -0.003 (-0.026, 0.020) | 0.778 |
| Caffeine Consumption | Parity | 75117 | 0.003 (-0.002, 0.008) | 0.262 | -0.005 (-0.010, 0.00060) | 0.079 |
| Caffeine Consumption | Sex Frequency | 73477 | -0.011 (-0.018, -0.004) | 0.001 | 0.018 ( 0.011, 0.024) | 3.36e-07 |
| Caffeine Consumption | Time to Conception | 55549 | 0.050 ( 0.026, 0.074) | 4.51e-05 | 0.008 (-0.017, 0.033) | 0.536 |
| BMI | Age at First Birth | 83128 | -0.042 (-0.049, -0.034) | 1.96e-29 | -0.017 (-0.023, -0.011) | 4.96e-08 |
| BMI | Parity | 83128 | -0.0009 (-0.002, 0.0005) | 0.193 | -0.004 (-0.005, -0.002) | 2.20e-07 |
| BMI | Sex Frequency | 81707 | -0.007 (-0.009, -0.005) | 5.87e-15 | -0.007 (-0.008, -0.005) | 1.37e-12 |
| BMI | Time to Conception | 61794 | 0.069 ( 0.063, 0.075) | 3.71e-99 | 0.061 ( 0.055, 0.068) | 2.04e-73 |
| Smoking Initiation | Age at First Birth | 82761 | -1.060 (-1.122, -0.998) | 1.79e-242 | -0.155 (-0.208, -0.103) | 5.68e-09 |
| Smoking Initiation | Parity | 82761 | -0.077 (-0.089, -0.065) | 7.20e-36 | -0.101 (-0.114, -0.089) | 3.71e-56 |
| Smoking Initiation | Sex Frequency | 80920 | 0.035 ( 0.019, 0.050) | 1.00e-05 | -0.020 (-0.036, -0.004) | 0.013 |
| Smoking Initiation | Time to Conception | 62567 | 0.133 ( 0.079, 0.187) | 1.30e-06 | 0.095 ( 0.038, 0.151) | 9.87e-04 |
| Smoking Heaviness | Age at First Birth | 16063 | -0.083 (-0.096, -0.070) | 2.07e-37 | -0.006 (-0.017, 0.005) | 0.313 |
| Smoking Heaviness | Parity | 16063 | -0.003 (-0.005, -0.00007) | 0.044 | -0.008 (-0.010, -0.005) | 4.25e-09 |
| Smoking Heaviness | Sex Frequency | 15671 | 0.013 ( 0.010, 0.017) | 1.17e-15 | 0.008 ( 0.005, 0.011) | 3.21e-06 |
| Smoking Heaviness | Time to Conception | 10223 | 0.032 ( 0.020, 0.045) | 1.82e-07 | 0.029 ( 0.016, 0.042) | 1.05e-05 |
| **Exposure** | **Outcome** | **N** | **OR (95% CI)** | **P-value** | **OR (95% CI)** | **P-value** |
| Alcohol Frequency | Subfertile | 60269 | 0.984 (0.960, 1.009) | 0.199 | 0.982 (0.957, 1.008) | 0.169 |
| Alcohol Frequency | Highly Fertile | 78165 | 1.023 (1.007, 1.038) | 0.004 | 1.021 (1.004, 1.037) | 0.013 |
| Alcohol Frequency | Infertility Treatment | 78595 | 0.900 (0.880, 0.920) | 5.75e-20 | 0.867 (0.847, 0.888) | 4.49e-32 |
| Alcohol Frequency | Miscarriage | 51637 | 0.996 (0.979, 1.014) | 0.668 | 0.986 (0.968, 1.004) | 0.118 |
| Binge Drinking | Subfertile | 59677 | 0.976 (0.947, 1.006) | 0.118 | 1.010 (0.979, 1.042) | 0.540 |
| Binge Drinking | Highly Fertile | 77272 | 1.027 (1.008, 1.046) | 0.004 | 1.008 (0.990, 1.028) | 0.383 |
| Binge Drinking | Infertility Treatment | 77674 | 0.761 (0.739, 0.783) | 8.82e-77 | 0.822 (0.798, 0.847) | 1.10e-37 |
| Binge Drinking | Miscarriage | 51029 | 0.951 (0.930, 0.972) | 6.98e-06 | 0.959 (0.938, 0.981) | 3.38e-04 |
| Caffeine Consumption | Subfertile | 55549 | 1.048 (1.024, 1.073) | 8.70e-05 | 1.010 (0.986, 1.034) | 0.433 |
| Caffeine Consumption | Highly Fertile | 73828 | 0.985 (0.971, 0.999) | 0.038 | 1.008 (0.994, 1.024) | 0.269 |
| Caffeine Consumption | Infertility Treatment | 73784 | 1.019 (0.998, 1.040) | 0.072 | 0.951 (0.931, 0.972) | 5.59e-06 |
| Caffeine Consumption | Miscarriage | 49315 | 1.011 (0.995, 1.027) | 0.192 | 1.001 (0.985, 1.018) | 0.905 |
| BMI | Subfertile | 61794 | 1.052 (1.047, 1.058) | 4.40e-72 | 1.048 (1.042, 1.054) | 2.84e-55 |
| BMI | Highly Fertile | 82089 | 0.983 (0.979, 0.987) | 2.07e-18 | 0.988 (0.984, 0.992) | 1.21e-09 |
| BMI | Infertility Treatment | 82045 | 1.037 (1.031, 1.042) | 5.16e-44 | 1.032 (1.026, 1.037) | 1.15e-29 |
| BMI | Miscarriage | 54353 | 1.007 (1.003, 1.012) | 5.46e-04 | 1.008 (1.004, 1.013) | 1.60e-04 |
| Smoking Initiation | Subfertile | 62567 | 1.108 (1.051, 1.168) | 1.28e-04 | 1.075 (1.018, 1.136) | 0.010 |
| Smoking Initiation | Highly Fertile | 81282 | 0.954 (0.923, 0.985) | 0.004 | 0.963 (0.931, 0.997) | 0.034 |
| Smoking Initiation | Infertility Treatment | 81743 | 0.935 (0.892, 0.981) | 0.006 | 0.965 (0.918, 1.015) | 0.165 |
| Smoking Initiation | Miscarriage | 53755 | 0.939 (0.905, 0.974) | 8.21e-04 | 0.963 (0.926, 1.001) | 0.055 |
| Smoking Heaviness | Subfertile | 10223 | 1.030 (1.020, 1.040) | 2.68e-09 | 1.029 (1.019, 1.040) | 4.89e-08 |
| Smoking Heaviness | Highly Fertile | 15704 | 0.998 (0.992, 1.005) | 0.571 | 1.002 (0.995, 1.009) | 0.534 |
| Smoking Heaviness | Infertility Treatment | 15674 | 1.004 (0.994, 1.013) | 0.467 | 1.005 (0.994, 1.015) | 0.393 |
| Smoking Heaviness | Miscarriage | 10488 | 1.005 (0.998, 1.012) | 0.196 | 1.002 (0.995, 1.010) | 0.531 |

**Supplementary Table S4. Observational associations between health behaviours in the fathers (full sample)**

|  |  |  | Unadjusted |  | Adjusted for birthyear and education | |
| --- | --- | --- | --- | --- | --- | --- |
| **Exposure** | **Outcome** | **N** | **Beta (95% CI)** | **P-value** | **Beta (95% CI)** | **P-value** |
| Alcohol Frequency | Age at First Birth | 63900 | 0.632 ( 0.598, 0.666) | 4.69e-288 | 0.329 ( 0.299, 0.359) | 4.25e-101 |
| Alcohol Frequency | Parity | 63900 | -0.052 (-0.058, -0.046) | 4.17e-63 | -0.056 (-0.063, -0.050) | 4.53e-65 |
| Alcohol Frequency | Sex Frequency | 61689 | 0.016 ( 0.008, 0.024) | 6.72e-05 | 0.042 ( 0.034, 0.050) | 7.01e-24 |
| Alcohol Frequency | Time to Conception | 49106 | 0.010 (-0.017, 0.037) | 0.473 | 0.023 (-0.007, 0.052) | 0.131 |
| Binge Drinking | Age at First Birth | 28637 | -0.125 (-0.187, -0.063) | 7.29e-05 | 0.261 ( 0.210, 0.311) | 5.01e-24 |
| Binge Drinking | Parity | 28637 | -0.083 (-0.094, -0.073) | 1.78e-56 | -0.073 (-0.084, -0.062) | 1.37e-39 |
| Binge Drinking | Sex Frequency | 27758 | 0.097 ( 0.083, 0.111) | 1.44e-42 | 0.076 ( 0.061, 0.090) | 4.52e-25 |
| Binge Drinking | Time to Conception | 22151 | -0.034 (-0.083, 0.015) | 0.178 | -0.006 (-0.057, 0.046) | 0.832 |
| Caffeine Consumption | Age at First Birth | 27445 | 0.204 ( 0.157, 0.252) | 4.38e-17 | -0.100 (-0.139, -0.061) | 5.13e-07 |
| Caffeine Consumption | Parity | 27445 | 0.017 ( 0.009, 0.025) | 3.18e-05 | 0.011 ( 0.003, 0.020) | 0.010 |
| Caffeine Consumption | Sex Frequency | 26585 | -0.024 (-0.035, -0.014) | 9.94e-06 | -0.004 (-0.016, 0.007) | 0.432 |
| Caffeine Consumption | Time to Conception | 21272 | 0.047 ( 0.010, 0.085) | 0.014 | 0.020 (-0.020, 0.059) | 0.332 |
| BMI | Age at First Birth | 67555 | 0.039 ( 0.028, 0.050) | 8.17e-12 | 0.026 ( 0.016, 0.035) | 2.23e-07 |
| BMI | Parity | 67555 | -0.003 (-0.005, -0.0008) | 0.006 | -0.006 (-0.008, -0.004) | 5.68e-09 |
| BMI | Sex Frequency | 65726 | -0.008 (-0.011, -0.006) | 2.94e-10 | -0.007 (-0.010, -0.005) | 2.92e-08 |
| BMI | Time to Conception | 52264 | 0.056 ( 0.047, 0.064) | 6.85e-34 | 0.045 ( 0.035, 0.054) | 2.30e-20 |
| Smoking Initiation | Age at First Birth | 65541 | -0.686 (-0.762, -0.610) | 5.45e-70 | -0.218 (-0.284, -0.151) | 1.55e-10 |
| Smoking Initiation | Parity | 65541 | -0.013 (-0.027, 0.0002) | 0.053 | -0.021 (-0.035, -0.007) | 0.004 |
| Smoking Initiation | Sex Frequency | 63215 | 0.007 (-0.011, 0.024) | 0.458 | -0.022 (-0.040, -0.004) | 0.018 |
| Smoking Initiation | Time to Conception | 50352 | 0.120 ( 0.060, 0.180) | 8.41e-05 | 0.041 (-0.023, 0.105) | 0.207 |
| Smoking Heaviness | Age at First Birth | 12570 | -0.039 (-0.054, -0.024) | 3.21e-07 | -0.009 (-0.022, 0.004) | 0.192 |
| Smoking Heaviness | Parity | 12570 | -0.003 (-0.006, -0.0005) | 0.020 | -0.004 (-0.007, -0.001) | 0.005 |
| Smoking Heaviness | Sex Frequency | 12014 | 0.009 ( 0.006, 0.012) | 6.88e-08 | 0.006 ( 0.002, 0.009) | 0.001 |
| Smoking Heaviness | Time to Conception | 8843 | 0.026 ( 0.014, 0.038) | 2.26e-05 | 0.021 ( 0.009, 0.034) | 0.001 |
| **Exposure** | **Outcome** | **N** | **OR (95% CI)** | **P-value** | **OR (95% CI)** | **P-value** |
| Alcohol Frequency | Subfertile | 49106 | 1.017 (0.990, 1.044) | 0.211 | 1.022 (0.993, 1.051) | 0.137 |
| Alcohol Frequency | Highly Fertile | 62476 | 1.015 (0.998, 1.032) | 0.076 | 1.009 (0.991, 1.027) | 0.337 |
| Binge Drinking | Subfertile | 22151 | 0.996 (0.949, 1.045) | 0.863 | 1.028 (0.977, 1.081) | 0.287 |
| Binge Drinking | Highly Fertile | 28077 | 1.044 (1.014, 1.076) | 0.004 | 1.026 (0.994, 1.059) | 0.107 |
| Caffeine Consumption | Subfertile | 21272 | 1.027 (0.989, 1.067) | 0.167 | 1.004 (0.965, 1.045) | 0.851 |
| Caffeine Consumption | Highly Fertile | 26911 | 0.981 (0.959, 1.004) | 0.106 | 0.997 (0.972, 1.021) | 0.780 |
| BMI | Subfertile | 52264 | 1.044 (1.036, 1.053) | 2.92e-24 | 1.037 (1.028, 1.046) | 2.09e-15 |
| BMI | Highly Fertile | 66548 | 0.976 (0.971, 0.982) | 9.10e-18 | 0.982 (0.976, 0.988) | 9.33e-10 |
| Smoking Initiation | Subfertile | 50352 | 1.079 (1.017, 1.144) | 0.011 | 1.003 (0.942, 1.068) | 0.915 |
| Smoking Initiation | Highly Fertile | 64056 | 0.958 (0.924, 0.994) | 0.023 | 0.984 (0.946, 1.023) | 0.408 |
| Smoking Heaviness | Subfertile | 8843 | 1.023 (1.013, 1.033) | 8.43e-06 | 1.019 (1.008, 1.030) | 5.68e-04 |
| Smoking Heaviness | Highly Fertile | 12254 | 0.999 (0.992, 1.005) | 0.668 | 1.000 (0.993, 1.007) | 0.973 |

**Supplementary Table S5. Observational associations between health behaviours in the mothers genotyped sample only**

|  |  |  | Unadjusted |  | Adjusted for birthyear and education | | Adjusted for birthyear, education and ADHD | |
| --- | --- | --- | --- | --- | --- | --- | --- | --- |
| **Exposure** | **Outcome** | **N** | **Beta (95% CI)** | **P-value** | **Beta (95% CI)** | **P-value** | **Beta (95% CI)** | **P-value** |
| Alcohol Frequency | Age at First Birth | 26815 | 0.890 ( 0.843, 0.937) | 1.12e-290 | 0.482 ( 0.443, 0.522) | 1.46e-126 | 0.498 ( 0.449, 0.548) | 1.70e-84 |
| Alcohol Frequency | Parity | 26815 | -0.093 (-0.102, -0.084) | 6.90e-88 | -0.093 (-0.103, -0.083) | 2.56e-80 | -0.087 (-0.099, -0.075) | 3.86e-44 |
| Alcohol Frequency | Sex Frequency | 26282 | 0.057 ( 0.045, 0.069) | 2.06e-20 | 0.086 ( 0.074, 0.099) | 3.18e-42 | 0.087 ( 0.071, 0.103) | 2.78e-27 |
| Alcohol Frequency | TTC | 20991 | -0.054 (-0.096, -0.012) | 0.011 | -0.051 (-0.095, -0.008) | 0.021 | -0.082 (-0.139, -0.025) | 0.005 |
| Binge Drinking | Age at First Birth | 26542 | -0.069 (-0.128, -0.010) | 0.021 | 0.329 ( 0.281, 0.377) | 1.31e-41 | 0.398 ( 0.337, 0.458) | 1.54e-37 |
| Binge Drinking | Parity | 26542 | -0.113 (-0.124, -0.102) | 5.15e-88 | -0.115 (-0.127, -0.104) | 3.00e-85 | -0.114 (-0.129, -0.100) | 3.85e-52 |
| Binge Drinking | Sex Frequency | 26020 | 0.111 ( 0.096, 0.125) | 3.09e-49 | 0.082 ( 0.067, 0.097) | 1.75e-26 | 0.083 ( 0.064, 0.102) | 1.69e-17 |
| Binge Drinking | TTC | 20795 | -0.025 (-0.076, 0.027) | 0.351 | 0.009 (-0.044, 0.062) | 0.742 | -0.021 (-0.090, 0.048) | 0.557 |
| Caffeine | Age at First Birth | 24508 | 0.355 ( 0.310, 0.401) | 5.08e-52 | 0.050 ( 0.012, 0.088) | 0.009 | 0.062 ( 0.013, 0.111) | 0.012 |
| Caffeine | Parity | 24508 | -0.007 (-0.016, 0.002) | 0.133 | -0.014 (-0.023, -0.004) | 0.003 | -0.013 (-0.024, -0.00090) | 0.035 |
| Caffeine | Sex Frequency | 24018 | -0.008 (-0.019, 0.004) | 0.183 | 0.017 ( 0.005, 0.029) | 0.004 | 0.015 (-0.000200, 0.030) | 0.054 |
| Caffeine | TTC | 19074 | 0.039 (-0.00070, 0.079) | 0.054 | 0.005 (-0.036, 0.047) | 0.795 | 0.009 (-0.045, 0.064) | 0.734 |
| BMI | Age at First Birth | 27268 | -0.049 (-0.062, -0.037) | 3.81e-15 | -0.018 (-0.028, -0.008) | 4.95e-04 | -0.008 (-0.021, 0.005) | 0.252 |
| BMI | Parity | 27268 | -0.002 (-0.005, 0.00003) | 0.053 | -0.004 (-0.006, -0.002) | 0.001 | -0.006 (-0.009, -0.002) | 8.50e-04 |
| BMI | Sex Frequency | 26858 | -0.006 (-0.009, -0.003) | 2.70e-04 | -0.007 (-0.010, -0.003) | 4.45e-05 | -0.008 (-0.013, -0.004) | 5.14e-05 |
| BMI | TTC | 21361 | 0.074 ( 0.063, 0.085) | 2.68e-41 | 0.066 ( 0.055, 0.077) | 9.37e-31 | 0.065 ( 0.050, 0.080) | 1.67e-17 |
| Smoking Initiation | Age at First Birth | 27597 | -0.896 (-1.000, -0.793) | 2.21e-64 | -0.024 (-0.110, 0.062) | 0.584 | 0.100 (-0.009, 0.210) | 0.072 |
| Smoking Initiation | Parity | 27597 | -0.079 (-0.099, -0.059) | 1.05e-14 | -0.105 (-0.126, -0.084) | 1.79e-22 | -0.122 (-0.149, -0.095) | 7.06e-19 |
| Smoking Initiation | Sex Frequency | 27038 | 0.045 ( 0.019, 0.071) | 7.86e-04 | -0.010 (-0.037, 0.017) | 0.449 | 0.008 (-0.026, 0.042) | 0.641 |
| Smoking Initiation | TTC | 21606 | 0.152 ( 0.062, 0.242) | 8.84e-04 | 0.088 (-0.005, 0.182) | 0.065 | 0.121 (-0.001, 0.243) | 0.053 |
| Smoking Heaviness | Age at First Birth | 4959 | -0.087 (-0.110, -0.065) | 3.19e-14 | -0.021 (-0.040, -0.001) | 0.037 | 0.029 ( 0.000900, 0.057) | 0.043 |
| Smoking Heaviness | Parity | 4959 | -0.002 (-0.006, 0.003) | 0.445 | -0.005 (-0.009, -0.00009) | 0.045 | -0.010 (-0.016, -0.004) | 0.002 |
| Smoking Heaviness | Sex Frequency | 4842 | 0.016 ( 0.010, 0.021) | 9.73e-08 | 0.012 ( 0.006, 0.018) | 1.01e-04 | 0.013 ( 0.005, 0.022) | 0.003 |
| Smoking Heaviness | TTC | 3411 | 0.038 ( 0.017, 0.058) | 4.33e-04 | 0.030 ( 0.008, 0.052) | 0.007 | 0.022 (-0.009, 0.053) | 0.161 |
| **Exposure** | **Outcome** | **N** | **OR (95% CI)** | **P-value** | **OR (95% CI)** | **P-value** | **OR (95% CI)** | **P-value** |
| Alcohol Frequency | Subfertile | 20991 | 0.966 (0.924, 1.009) | 0.117 | 0.963 (0.920, 1.008) | 0.106 | 0.917 (0.865, 0.973) | 0.004 |
| Alcohol Frequency | Highly Fertile | 26407 | 1.033 (1.006, 1.060) | 0.015 | 1.030 (1.002, 1.058) | 0.036 | 1.040 (1.004, 1.077) | 0.029 |
| Alcohol Frequency | Infertility Treatment | 26538 | 0.878 (0.843, 0.914) | 3.15e-10 | 0.849 (0.814, 0.886) | 3.45e-14 | 0.819 (0.776, 0.864) | 5.00e-13 |
| Alcohol Frequency | Miscarriage | 17139 | 0.989 (0.960, 1.020) | 0.488 | 0.981 (0.950, 1.012) | 0.226 | 0.985 (0.945, 1.026) | 0.460 |
| Binge Drinking | Subfertile | 20795 | 0.985 (0.933, 1.039) | 0.581 | 1.017 (0.962, 1.075) | 0.563 | 0.978 (0.911, 1.050) | 0.542 |
| Binge Drinking | Highly Fertile | 26142 | 1.039 (1.007, 1.072) | 0.017 | 1.022 (0.989, 1.057) | 0.186 | 1.021 (0.978, 1.065) | 0.344 |
| Binge Drinking | Infertility Treatment | 26267 | 0.742 (0.705, 0.782) | 4.48e-29 | 0.796 (0.755, 0.840) | 1.12e-16 | 0.760 (0.709, 0.815) | 1.17e-14 |
| Binge Drinking | Miscarriage | 16978 | 0.964 (0.927, 1.002) | 0.065 | 0.965 (0.928, 1.005) | 0.084 | 0.973 (0.924, 1.026) | 0.315 |
| Caffeine Consumption | Subfertile | 19074 | 1.041 (0.999, 1.085) | 0.059 | 1.006 (0.964, 1.050) | 0.796 | 1.014 (0.960, 1.071) | 0.615 |
| Caffeine Consumption | Highly Fertile | 24142 | 0.992 (0.968, 1.017) | 0.511 | 1.007 (0.982, 1.034) | 0.579 | 1.018 (0.984, 1.053) | 0.305 |
| Caffeine Consumption | Infertility Treatment | 24216 | 1.008 (0.971, 1.046) | 0.680 | 0.943 (0.908, 0.980) | 0.003 | 0.954 (0.908, 1.002) | 0.062 |
| Caffeine Consumption | Miscarriage | 15780 | 1.009 (0.981, 1.038) | 0.539 | 1.003 (0.974, 1.033) | 0.831 | 1.006 (0.968, 1.046) | 0.747 |
| BMI | Subfertile | 21361 | 1.055 (1.045, 1.066) | 6.31e-26 | 1.049 (1.038, 1.060) | 3.30e-19 | 1.056 (1.042, 1.071) | 4.39e-15 |
| BMI | Highly Fertile | 26991 | 0.978 (0.972, 0.985) | 4.05e-10 | 0.982 (0.975, 0.989) | 6.39e-07 | 0.982 (0.972, 0.991) | 1.43e-04 |
| BMI | Infertility Treatment | 27084 | 1.040 (1.030, 1.049) | 4.55e-17 | 1.033 (1.023, 1.043) | 5.55e-11 | 1.036 (1.023, 1.050) | 3.61e-08 |
| BMI | Miscarriage | 17531 | 1.010 (1.003, 1.018) | 0.006 | 1.011 (1.003, 1.019) | 0.006 | 1.015 (1.004, 1.025) | 0.006 |
| Smoking Initiation | Subfertile | 21606 | 1.133 (1.032, 1.243) | 0.009 | 1.086 (0.985, 1.197) | 0.099 | 1.074 (0.948, 1.217) | 0.265 |
| Smoking Initiation | Highly Fertile | 27174 | 0.930 (0.880, 0.983) | 0.011 | 0.949 (0.895, 1.007) | 0.085 | 0.903 (0.836, 0.975) | 0.009 |
| Smoking Initiation | Infertility Treatment | 27315 | 0.952 (0.875, 1.036) | 0.256 | 0.955 (0.873, 1.044) | 0.309 | 0.951 (0.848, 1.067) | 0.395 |
| Smoking Initiation | Miscarriage | 17678 | 0.984 (0.923, 1.050) | 0.629 | 1.000 (0.934, 1.070) | 0.996 | 1.030 (0.942, 1.126) | 0.516 |
| Smoking Heaviness | Subfertile | 3411 | 1.032 (1.014, 1.049) | 2.96e-04 | 1.029 (1.011, 1.048) | 0.002 | 1.027 (1.002, 1.053) | 0.037 |
| Smoking Heaviness | Highly Fertile | 4858 | 0.991 (0.980, 1.003) | 0.145 | 0.997 (0.985, 1.010) | 0.652 | 1.002 (0.984, 1.020) | 0.834 |
| Smoking Heaviness | Infertility Treatment | 4880 | 1.000 (0.982, 1.019) | 0.974 | 0.997 (0.977, 1.016) | 0.727 | 1.002 (0.975, 1.029) | 0.905 |
| Smoking Heaviness | Miscarriage | 3174 | 1.008 (0.995, 1.021) | 0.255 | 1.008 (0.995, 1.022) | 0.228 | 0.996 (0.976, 1.016) | 0.693 |

**Supplementary Table S6. Observational associations between health behaviours in the fathers genotyped sample only**

|  |  |  | Unadjusted |  | Adjusted for birthyear and education | | Adjusted for birthyear, education and ADHD | |
| --- | --- | --- | --- | --- | --- | --- | --- | --- |
| **Exposure** | **Outcome** | **N** | **Beta (95% CI)** | **P-value** | **Beta (95% CI)** | **P-value** |  |  |
| Alcohol Frequency | Age at First Birth | 24763 | 0.667 ( 0.613, 0.722) | 6.36e-128 | 0.345 ( 0.297, 0.392) | 9.13e-46 | 0.266 ( 0.200, 0.332) | 3.48e-15 |
| Alcohol Frequency | Parity | 24763 | -0.050 (-0.059, -0.040) | 6.89e-24 | -0.054 (-0.064, -0.044) | 1.30e-24 | -0.048 (-0.063, -0.034) | 5.39e-11 |
| Alcohol Frequency | Sex Frequency | 24057 | 0.026 ( 0.014, 0.039) | 3.50e-05 | 0.055 ( 0.042, 0.068) | 4.61e-16 | 0.065 ( 0.046, 0.084) | 3.42e-11 |
| Alcohol Frequency | Time to Conception | 19497 | 0.010 (-0.034, 0.053) | 0.661 | 0.009 (-0.038, 0.055) | 0.717 | -0.009 (-0.075, 0.058) | 0.800 |
| Binge Drinking | Age at First Birth | 10755 | -0.027 (-0.127, 0.073) | 0.596 | 0.326 ( 0.246, 0.406) | 1.88e-15 | 0.332 ( 0.251, 0.412) | 1.01e-15 |
| Binge Drinking | Parity | 10755 | -0.093 (-0.110, -0.076) | 1.24e-27 | -0.080 (-0.097, -0.063) | 2.43e-19 | -0.082 (-0.099, -0.064) | 9.96e-20 |
| Binge Drinking | Sex Frequency | 10464 | 0.091 ( 0.069, 0.114) | 4.49e-15 | 0.071 ( 0.048, 0.094) | 2.78e-09 | 0.074 ( 0.051, 0.098) | 6.96e-10 |
| Binge Drinking | Time to Conception | 8471 | -0.035 (-0.114, 0.044) | 0.383 | -0.003 (-0.085, 0.078) | 0.933 | -0.002 (-0.084, 0.080) | 0.955 |
| Caffeine Consumption | Age at First Birth | 10320 | 0.192 ( 0.118, 0.267) | 4.73e-07 | -0.105 (-0.166, -0.044) | 7.03e-04 | -0.109 (-0.170, -0.048) | 4.67e-04 |
| Caffeine Consumption | Parity | 10320 | 0.014 ( 0.001, 0.027) | 0.032 | 0.014 ( 0.00050, 0.027) | 0.042 | 0.013 (-0.00040, 0.026) | 0.057 |
| Caffeine Consumption | Sex Frequency | 10043 | -0.034 (-0.051, -0.017) | 9.73e-05 | -0.016 (-0.033, 0.002) | 0.086 | -0.014 (-0.031, 0.004) | 0.139 |
| Caffeine Consumption | Time to Conception | 8146 | 0.018 (-0.040, 0.076) | 0.538 | -0.002 (-0.063, 0.058) | 0.942 | 0.002 (-0.059, 0.063) | 0.939 |
| BMI | Age at First Birth | 26149 | 0.034 ( 0.016, 0.051) | 1.53e-04 | 0.020 ( 0.005, 0.035) | 0.009 | -0.005 (-0.026, 0.017) | 0.666 |
| BMI | Parity | 26149 | -0.004 (-0.008, -0.001) | 0.006 | -0.007 (-0.011, -0.004) | 1.02e-05 | -0.008 (-0.013, -0.003) | 7.38e-04 |
| BMI | Sex Frequency | 25526 | -0.008 (-0.012, -0.004) | 3.76e-05 | -0.008 (-0.012, -0.004) | 1.14e-04 | -0.003 (-0.009, 0.003) | 0.324 |
| BMI | Time to Conception | 20654 | 0.049 ( 0.035, 0.063) | 5.89e-12 | 0.038 ( 0.023, 0.053) | 4.09e-07 | 0.044 ( 0.023, 0.066) | 5.51e-05 |
| Smoking Initiation | Age at First Birth | 25312 | -0.564 (-0.682, -0.445) | 1.07e-20 | -0.160 (-0.262, -0.058) | 0.002 | -0.019 (-0.167, 0.129) | 0.803 |
| Smoking Initiation | Parity | 25312 | -0.036 (-0.057, -0.015) | 7.88e-04 | -0.041 (-0.063, -0.018) | 3.44e-04 | -0.048 (-0.080, -0.016) | 0.004 |
| Smoking Initiation | Sex Frequency | 24579 | -0.006 (-0.033, 0.022) | 0.690 | -0.027 (-0.055, 0.002) | 0.066 | -0.015 (-0.058, 0.028) | 0.500 |
| Smoking Initiation | Time to Conception | 19937 | 0.115 ( 0.022, 0.208) | 0.016 | 0.040 (-0.059, 0.139) | 0.426 | -0.022 (-0.169, 0.125) | 0.771 |
| Smoking Heaviness | Age at First Birth | 4715 | -0.027 (-0.051, -0.002) | 0.032 | 0.003 (-0.018, 0.025) | 0.758 | 0.009 (-0.024, 0.042) | 0.578 |
| Smoking Heaviness | Parity | 4715 | -0.005 (-0.009, -0.00040) | 0.034 | -0.006 (-0.010, -0.001) | 0.014 | -0.009 (-0.016, -0.002) | 0.012 |
| Smoking Heaviness | Sex Frequency | 4547 | 0.009 ( 0.004, 0.015) | 0.001 | 0.007 ( 0.00080, 0.012) | 0.026 | 0.005 (-0.004, 0.014) | 0.287 |
| Smoking Heaviness | Time to Conception | 3450 | 0.021 ( 0.002, 0.041) | 0.031 | 0.015 (-0.005, 0.036) | 0.144 | -0.009 (-0.043, 0.025) | 0.597 |
| **Exposure** | **Outcome** | **N** | **OR (95% CI)** | **P-value** | **OR (95% CI)** | **P-value** | **OR (95% CI)** | **P-value** |
| Alcohol Frequency | Subfertile | 19497 | 1.018 (0.973, 1.064) | 0.438 | 1.010 (0.963, 1.059) | 0.688 | 1.006 (0.937, 1.080) | 0.866 |
| Alcohol Frequency | Highly Fertile | 24319 | 1.011 (0.985, 1.039) | 0.409 | 1.017 (0.988, 1.047) | 0.258 | 1.046 (1.002, 1.092) | 0.039 |
| Binge Drinking | Subfertile | 8471 | 1.024 (0.941, 1.113) | 0.586 | 1.066 (0.977, 1.164) | 0.149 | 1.066 (0.976, 1.163) | 0.156 |
| Binge Drinking | Highly Fertile | 10568 | 1.046 (0.996, 1.099) | 0.071 | 1.042 (0.989, 1.097) | 0.121 | 1.045 (0.992, 1.101) | 0.096 |
| Caffeine Consumption | Subfertile | 8146 | 1.031 (0.967, 1.099) | 0.353 | 1.013 (0.946, 1.085) | 0.710 | 1.014 (0.946, 1.086) | 0.700 |
| Caffeine Consumption | Highly Fertile | 10144 | 1.010 (0.973, 1.049) | 0.589 | 1.026 (0.986, 1.068) | 0.202 | 1.028 (0.987, 1.071) | 0.177 |
| BMI | Subfertile | 20654 | 1.045 (1.031, 1.060) | 3.52e-10 | 1.038 (1.023, 1.053) | 7.83e-07 | 1.048 (1.025, 1.071) | 2.80e-05 |
| BMI | Highly Fertile | 25813 | 0.980 (0.971, 0.988) | 4.53e-06 | 0.983 (0.974, 0.992) | 3.16e-04 | 0.987 (0.973, 1.001) | 0.061 |
| Smoking Initiation | Subfertile | 19937 | 1.111 (1.008, 1.223) | 0.033 | 1.031 (0.930, 1.142) | 0.566 | 1.014 (0.864, 1.190) | 0.863 |
| Smoking Initiation | Highly Fertile | 24854 | 0.978 (0.923, 1.037) | 0.461 | 1.001 (0.941, 1.066) | 0.966 | 1.061 (0.964, 1.168) | 0.223 |
| Smoking Heaviness | Subfertile | 3450 | 1.020 (1.003, 1.037) | 0.021 | 1.013 (0.995, 1.031) | 0.168 | 0.999 (0.969, 1.030) | 0.966 |
| Smoking Heaviness | Highly Fertile | 4630 | 0.997 (0.985, 1.008) | 0.564 | 1.000 (0.988, 1.012) | 0.953 | 1.002 (0.982, 1.021) | 0.879 |

**Supplementary Table S7. Individual-level Mendelian randomisation analysis in the mothers**

| **Exposure** | **Outcome** | **N** | **Beta/RD (95% CI)** | **P-value** |
| --- | --- | --- | --- | --- |
| Alcohol Consumption | Age at First Birth | 26,799 | 0.536 (-0.655, 1.728) | 0.378 |
|  | Parity | 26,799 | -0.133 (-0.408, 0.142) | 0.342 |
|  | Sex Frequency | 26,265 | 0.113 (-0.259, 0.485) | 0.550 |
|  | Time to Conception | 20,974 | 0.652 (-0.807, 2.111) | 0.381 |
|  | Subfertile | 20,974 | 0.058 (-0.067, 0.183) | 0.366 |
|  | Highly Fertile | 26,390 | -0.097 (-0.247, 0.052) | 0.201 |
|  | Infertility Treatment | 26,521 | -0.022 (-0.114, 0.071) | 0.649 |
|  | Miscarriage | 17,122 | 0.020 (-0.222, 0.262) | 0.871 |
| Caffeine Consumption | Age at First Birth | 24,462 | 0.731 (-0.211, 1.673) | 0.128 |
|  | Parity | 24,462 | 0.054 (-0.155, 0.263) | 0.612 |
|  | Sex Frequency | 23,975 | -0.116 (-0.398, 0.166) | 0.420 |
|  | Time to Conception | 19,057 | -0.214 (-1.284, 0.856) | 0.695 |
|  | Subfertile | 19,057 | 0.005 (-0.087, 0.096) | 0.919 |
|  | Highly Fertile | 24,095 | -0.014 (-0.122, 0.094) | 0.804 |
|  | Infertility Treatment | 24,175 | 0.082 ( 0.006, 0.159) | 0.034 |
|  | Miscarriage | 15,737 | -0.009 (-0.164, 0.146) | 0.908 |
| BMI | Age at First Birth | 27,216 | -0.113 (-0.153, -0.073) | 2.82e-08 |
|  | Parity | 27,216 | -0.0004 (-0.010, 0.009) | 0.940 |
|  | Sex Frequency | 26,810 | 0.009 (-0.003, 0.021) | 0.132 |
|  | Time to Conception | 21,344 | 0.028 (-0.015, 0.070) | 0.205 |
|  | Subfertile | 21,344 | 0.005 ( 0.001, 0.008) | 0.011 |
|  | Highly Fertile | 26,938 | 0.002 (-0.002, 0.007) | 0.325 |
|  | Infertility Treatment | 27,038 | 0.004 ( 0.0006, 0.007) | 0.020 |
|  | Miscarriage | 17,484 | 0.0002 (-0.006, 0.006) | 0.947 |
| Smoking Initiation | Age at First Birth | 27,581 | -2.647 (-3.565, -1.730) | 1.57e-08 |
|  | Parity | 27,581 | -0.111 (-0.315, 0.093) | 0.286 |
|  | Sex Frequency | 27,021 | 0.126 (-0.138, 0.390) | 0.349 |
|  | Time to Conception | 21,589 | -0.412 (-1.281, 0.458) | 0.354 |
|  | Subfertile | 21,589 | -0.037 (-0.112, 0.037) | 0.329 |
|  | Highly Fertile | 27,157 | 0.009 (-0.096, 0.113) | 0.873 |
|  | Infertility Treatment | 27,298 | 0.012 (-0.056, 0.080) | 0.730 |
|  | Miscarriage | 17,661 | -0.113 (-0.255, 0.028) | 0.117 |
| Smoking Heaviness | Age at First Birth | 4,889 | 0.040 (-0.135, 0.215) | 0.657 |
|  | Parity | 4,889 | 0.017 (-0.021, 0.056) | 0.378 |
|  | Sex Frequency | 4,774 | -0.025 (-0.078, 0.028) | 0.362 |
|  | Time to Conception | 3,390 | 0.034 (-0.165, 0.234) | 0.738 |
|  | Subfertile | 3,390 | -0.0009 (-0.019, 0.017) | 0.923 |
|  | Highly Fertile | 4,791 | -0.009 (-0.027, 0.009) | 0.338 |
|  | Infertility Treatment | 4,822 | 0.002 (-0.009, 0.014) | 0.682 |
|  | Miscarriage | 3,125 | -0.020 (-0.044, 0.004) | 0.104 |

**Supplementary Table S8. Individual-level Mendelian randomisation analysis in the fathers**

| **Exposure** | **Outcome** | **N** | **Beta/RD (95% CI)** | **P-value** |
| --- | --- | --- | --- | --- |
| Alcohol Consumption | Age at First Birth | 24,746 | -1.005 (-2.232, 0.223) | 0.109 |
|  | Parity | 24,746 | 0.165 (-0.088, 0.417) | 0.201 |
|  | Sex Frequency | 24,040 | 0.046 (-0.292, 0.384) | 0.790 |
|  | Time to Conception | 19,480 | 0.439 (-0.609, 1.486) | 0.412 |
|  | Subfertile | 19,480 | 0.020 (-0.069, 0.109) | 0.659 |
|  | Highly Fertile | 24,301 | -0.016 (-0.146, 0.114) | 0.808 |
| Caffeine Consumption | Age at First Birth | 10,303 | -0.264 (-1.401, 0.874) | 0.650 |
|  | Parity | 10,303 | 0.028 (-0.217, 0.273) | 0.824 |
|  | Sex Frequency | 10,027 | -0.106 (-0.436, 0.225) | 0.532 |
|  | Time to Conception | 8,130 | -0.149 (-1.164, 0.865) | 0.773 |
|  | Subfertile | 8,130 | -0.072 (-0.163, 0.019) | 0.121 |
|  | Highly Fertile | 10,127 | -0.043 (-0.177, 0.090) | 0.526 |
| BMI | Age at First Birth | 26,131 | -0.171 (-0.232, -0.111) | 2.44e-08 |
|  | Parity | 26,131 | -0.0008 (-0.013, 0.012) | 0.906 |
|  | Sex Frequency | 25,506 | 0.002 (-0.014, 0.019) | 0.772 |
|  | Time to Conception | 20,636 | 0.018 (-0.039, 0.075) | 0.529 |
|  | Subfertile | 20,636 | 0.002 (-0.003, 0.007) | 0.445 |
|  | Highly Fertile | 25,792 | 0.0008 (-0.006, 0.007) | 0.799 |
| Smoking Initiation | Age at First Birth | 25,295 | -2.824 (-4.072, -1.575) | 9.36e-06 |
|  | Parity | 25,295 | -0.016 (-0.272, 0.241) | 0.904 |
|  | Sex Frequency | 24,562 | -0.107 (-0.440, 0.227) | 0.531 |
|  | Time to Conception | 19,920 | 1.468 ( 0.286, 2.650) | 0.015 |
|  | Subfertile | 19,920 | 0.029 (-0.070, 0.129) | 0.565 |
|  | Highly Fertile | 24,836 | -0.081 (-0.214, 0.051) | 0.229 |
| Smoking Heaviness | Age at First Birth | 4,690 | 0.012 (-0.203, 0.227) | 0.915 |
|  | Parity | 4,690 | -0.056 (-0.103, -0.009) | 0.019 |
|  | Sex Frequency | 4,523 | 0.033 (-0.027, 0.093) | 0.277 |
|  | Time to Conception | 3,426 | 0.117 (-0.083, 0.316) | 0.253 |
|  | Subfertile | 3,426 | 0.019 ( 0.0006, 0.037) | 0.043 |
|  | Highly Fertile | 4,605 | -0.014 (-0.035, 0.007) | 0.189 |

**Supplementary Table S9. Evidence for assortative mating: the association of PGS on partners health behaviours and correlation between PGS**

| **PGS** | **Exposure in Partner** | **N** | **Beta/OR (95% CI)** | **P-value** |
| --- | --- | --- | --- | --- |
| Mother DPW p<5x10-8 | Father Alcohol Frequency | 24,604 | 0.010 (-0.003, 0.024) | 0.144 |
| Father DPW p<5x10-8 | Mother Alcohol Frequency | 25,882 | 0.016 ( 0.003, 0.029) | 0.018 |
| Mother Caffeine p<5x10-6 | Father Caffeine Consumption | 10,393 | 0.026 ( 0.002, 0.049) | 0.034 |
| Father Caffeine p<5x10-6 | Mother Caffeine Consumption | 23,530 | 0.008 (-0.007, 0.024) | 0.290 |
| Mother BMI p<5x10-8 | Father BMI | 27,200 | 0.164 ( 0.124, 0.203) | 4.18e-16 |
| Father BMI p<5x10-8 | Mother BMI | 26,172 | 0.177 ( 0.126, 0.229) | 1.16e-11 |
| Mother SI p<5x10-8 | Father Smoking Initiation | 25,185 | 1.067 ( 1.041, 1.094) | 3.48e-07 |
| Father SI p<5x10-8 | Mother Smoking Initiation | 26,635 | 1.072 ( 1.046, 1.098) | 2.01e-08 |
| Mother CPD p<5x10-8 | Father Smoking Heaviness | 4,755 | 0.050 (-0.126, 0.226) | 0.578 |
| Father CPD p<5x10-8 | Mother Smoking Heaviness | 4,648 | 0.054 (-0.112, 0.220) | 0.524 |
| **PGS** | **PGS** | **N** | **r** | **P-value** |
| Mother DPW p<5x10-8 | Father DPW p<5x10-8 | 22,534 | 0.017 | 0.01 |
| Mother Caffeine p<5x10-6 | Father Caffeine p<5x10-6 | 22,534 | 0.003 | 0.60 |
| Mother BMI p<5x10-8 | Father BMI p<5x10-8 | 22,534 | 0.016 | 0.01 |
| Mother SI p<5x10-8 | Father SI p<5x10-8 | 22,534 | 0.013 | 0.05 |
| Mother CPD p<5x10-8 | Father CPD p<5x10-8 | 22,534 | 0.002 | 0.79 |

**Supplementary Table S10. Evidence for reintroduced confounding: associations between PGS and other health behaviours or confounders**

| **PRS** | **Confounder** | **N** | **Beta (95% CI)** | **P-value** |
| --- | --- | --- | --- | --- |
| Mother DPW p<5x10-8 | Age | 28,839 | -0.067 (-0.119, -0.015) | 0.011 |
| Mother DPW p<5x10-8 | Income | 26,307 | -0.030 (-0.056, -0.003) | 0.028 |
| Mother DPW p<5x10-8 | Binge Drinking | 26,544 | 0.031 ( 0.020, 0.042) | 1.53e-08 |
| Mother DPW p<5x10-8 | Caffeine Consumption | 24,510 | 0.019 ( 0.004, 0.035) | 0.012 |
| Mother DPW p<5x10-8 | BMI | 27,270 | -0.007 (-0.057, 0.044) | 0.796 |
| Mother DPW p<5x10-8 | Smoking Heaviness | 4,959 | 0.004 (-0.161, 0.169) | 0.961 |
| Mother Caffeine p<5x10-8 | Age | 28,839 | -0.055 (-0.107, -0.003) | 0.038 |
| Mother Caffeine p<5x10-8 | Income | 26,307 | -0.002 (-0.028, 0.025) | 0.895 |
| Mother Caffeine p<5x10-8 | Alcohol Frequency | 26,817 | 0.001 (-0.012, 0.014) | 0.840 |
| Mother Caffeine p<5x10-8 | Binge Drinking | 26,544 | 0.002 (-0.009, 0.013) | 0.682 |
| Mother Caffeine p<5x10-8 | BMI | 27,270 | -0.052 (-0.103, -0.002) | 0.043 |
| Mother Caffeine p<5x10-8 | Smoking Heaviness | 4,959 | -0.009 (-0.173, 0.154) | 0.910 |
| Mother BMI p<5x10-8 | Age | 28,839 | -0.131 (-0.183, -0.079) | 7.98e-07 |
| Mother BMI p<5x10-8 | Income | 26,307 | -0.076 (-0.102, -0.049) | 2.05e-08 |
| Mother BMI p<5x10-8 | Alcohol Frequency | 26,817 | -0.031 (-0.045, -0.018) | 2.50e-06 |
| Mother BMI p<5x10-8 | Binge Drinking | 26,544 | 0.009 (-0.001, 0.020) | 0.088 |
| Mother BMI p<5x10-8 | Caffeine Consumption | 24,510 | 0.039 ( 0.024, 0.054) | 5.08e-07 |
| Mother BMI p<5x10-8 | Smoking Heaviness | 4,959 | 0.315 ( 0.152, 0.478) | 1.52e-04 |
| Mother SI p<5x10-8 | Age | 28,839 | -0.117 (-0.169, -0.066) | 9.30e-06 |
| Mother SI p<5x10-8 | Income | 26,307 | -0.081 (-0.107, -0.054) | 2.61e-09 |
| Mother SI p<5x10-8 | Alcohol Frequency | 26,817 | 0.0009 (-0.012, 0.014) | 0.894 |
| Mother SI p<5x10-8 | Binge Drinking | 26,544 | 0.040 ( 0.029, 0.051) | 7.39e-13 |
| Mother SI p<5x10-8 | Caffeine Consumption | 24,510 | 0.063 ( 0.048, 0.078) | 4.05e-16 |
| Mother SI p<5x10-8 | BMI | 27,270 | 0.138 ( 0.087, 0.188) | 9.54e-08 |
| Mother SI p<5x10-8 | Smoking Heaviness | 4,959 | 0.233 ( 0.068, 0.398) | 0.006 |
| Mother CPD p<5x10-8 | Age | 28,839 | -0.015 (-0.067, 0.037) | 0.568 |
| Mother CPD p<5x10-8 | Income | 26,307 | -0.013 (-0.040, 0.013) | 0.324 |
| Mother CPD p<5x10-8 | Alcohol Frequency | 26,817 | -0.024 (-0.037, -0.011) | 2.35e-04 |
| Mother CPD p<5x10-8 | Binge Drinking | 26,544 | -0.008 (-0.019, 0.003) | 0.151 |
| Mother CPD p<5x10-8 | Caffeine Consumption | 24,510 | 0.012 (-0.003, 0.027) | 0.116 |
| Mother CPD p<5x10-8 | BMI | 27,270 | 0.048 (-0.002, 0.098) | 0.061 |
| Father DPW p<5x10-8 | Age | 28,839 | -0.067 (-0.119, -0.015) | 0.011 |
| Father DPW p<5x10-8 | Income | 26,307 | -0.030 (-0.056, -0.003) | 0.028 |
| Father DPW p<5x10-8 | Binge Drinking | 26,544 | 0.031 ( 0.020, 0.042) | 1.53e-08 |
| Father DPW p<5x10-8 | Caffeine Consumption | 24,510 | 0.019 ( 0.004, 0.035) | 0.012 |
| Father DPW p<5x10-8 | BMI | 27,270 | -0.007 (-0.057, 0.044) | 0.796 |
| Father DPW p<5x10-8 | Smoking Heaviness | 4,959 | 0.004 (-0.161, 0.169) | 0.961 |
| Father Caffeine p<5x10-8 | Age | 28,839 | -0.055 (-0.107, -0.003) | 0.038 |
| Father Caffeine p<5x10-8 | Income | 26,307 | -0.002 (-0.028, 0.025) | 0.895 |
| Father Caffeine p<5x10-8 | Alcohol Frequency | 26,817 | 0.001 (-0.012, 0.014) | 0.840 |
| Father Caffeine p<5x10-8 | Binge Drinking | 26,544 | 0.002 (-0.009, 0.013) | 0.682 |
| Father Caffeine p<5x10-8 | BMI | 27,270 | -0.052 (-0.103, -0.002) | 0.043 |
| Father Caffeine p<5x10-8 | Smoking Heaviness | 4,959 | -0.009 (-0.173, 0.154) | 0.910 |
| Father BMI p<5x10-8 | Age | 28,839 | -0.131 (-0.183, -0.079) | 7.98e-07 |
| Father BMI p<5x10-8 | Income | 26,307 | -0.076 (-0.102, -0.049) | 2.05e-08 |
| Father BMI p<5x10-8 | Alcohol Frequency | 26,817 | -0.031 (-0.045, -0.018) | 2.50e-06 |
| Father BMI p<5x10-8 | Binge Drinking | 26,544 | 0.009 (-0.001, 0.020) | 0.088 |
| Father BMI p<5x10-8 | Caffeine Consumption | 24,510 | 0.039 ( 0.024, 0.054) | 5.08e-07 |
| Father BMI p<5x10-8 | Smoking Heaviness | 4,959 | 0.315 ( 0.152, 0.478) | 1.52e-04 |
| Father SI p<5x10-8 | Age | 28,839 | -0.117 (-0.169, -0.066) | 9.30e-06 |
| Father SI p<5x10-8 | Income | 26,307 | -0.081 (-0.107, -0.054) | 2.61e-09 |
| Father SI p<5x10-8 | Alcohol Frequency | 26,817 | 0.0009 (-0.012, 0.014) | 0.894 |
| Father SI p<5x10-8 | Binge Drinking | 26,544 | 0.040 ( 0.029, 0.051) | 7.39e-13 |
| Father SI p<5x10-8 | Caffeine Consumption | 24,510 | 0.063 ( 0.048, 0.078) | 4.05e-16 |
| Father SI p<5x10-8 | BMI | 27,270 | 0.138 ( 0.087, 0.188) | 9.54e-08 |
| Father SI p<5x10-8 | Smoking Heaviness | 4,959 | 0.233 ( 0.068, 0.398) | 0.006 |
| Father CPD p<5x10-8 | Age | 28,839 | -0.015 (-0.067, 0.037) | 0.568 |
| Father CPD p<5x10-8 | Income | 26,307 | -0.013 (-0.040, 0.013) | 0.324 |
| Father CPD p<5x10-8 | Alcohol Frequency | 26,817 | -0.024 (-0.037, -0.011) | 2.35e-04 |
| Father CPD p<5x10-8 | Binge Drinking | 26,544 | -0.008 (-0.019, 0.003) | 0.151 |
| Father CPD p<5x10-8 | Caffeine Consumption | 24,510 | 0.012 (-0.003, 0.027) | 0.116 |
| Father CPD p<5x10-8 | BMI | 27,270 | 0.048 (-0.002, 0.098) | 0.061 |

**Supplementary Table S11. Summary level MR sensitivity tests conducted in the MoBa sample**

| Exposure | Outcome | Method | Beta (95% CI) | P-value |
| --- | --- | --- | --- | --- |
| BMI | Age at first birth | IVW | -0.450 (-0.568, -0.331) | 1.06E-13 |
|  |  | MR Egger | -0.356 (-0.689, -0.023) | 0.04 |
|  |  | *MR Egger Intercept* | *-0.001 (-0.006, 0.003)* | *0.56* |
|  |  | Weighted Median | -0.350 (-0.547, -0.152) | 0.0005 |
|  |  | Weighted Mode | -0.119 (-0.644, 0.407) | 0.66 |
| Smoking Initiation | Age at first birth | IVW | -0.697 (-0.917, -0.477) | 5.54E-10 |
|  |  | MR Egger | -0.844 (-1.770, 0.081) | 0.08 |
|  |  | *MR Egger Intercept* | *0.003 (-0.015, 0.021)* | *0.75* |
|  |  | Weighted Median | -0.713 (-0.995, -0.431) | 7.31E-07 |
|  |  | Weighted Mode | -1.084 (-1.997, -0.171) | 0.02 |

**Supplementary Table S12. Summary of mother characteristics comparing planners and non-planners**

|  | Non-Planners | Planners |  |
| --- | --- | --- | --- |
|  | **Mean /%** | **Mean /%** | **P-value** |
| Age (years) | 30.62 | 30.70 | 0.78 |
| University education | 47% | 67% | <0.001 |
| BMI (kg/m2) | 23.97 | 24.07 | 0.009 |
| Alcohol consumption |  |  | <0.001 |
| *Never* | 7.6% | 7.0% |  |
| *Daily* | 0.3% | 0.2% |  |
| Binge drinking |  |  | <0.001 |
| *Never* | 29% | 34% |  |
| *Several times per week* | 1.8% | 0.6% |  |
| Ever smokers | 59% | 48% | <0.001 |
| Smoking heaviness (cigarettes per day) | 12.37 | 10.97 | <0.001 |
| Caffeine consumption (mg per day) | 148.22 | 139.76 | <0.001 |
| Age at first birth (years) | 25.84 | 27.65 | <0.001 |
| Number of children (N children) | 2.63 | 2.51 | <0.001 |

**Supplementary Table S13. Sensitivity analysis in mothers including non-planners in time to conception observational analyses compared with primary analysis with planners only**

|  | **Planners only** | | | **Including non-planners** | | |
| --- | --- | --- | --- | --- | --- | --- |
| **Exposure** | **N** | **Beta (95% CI)** | **P-value** | **N** | **Beta (95% CI)** | **P-value** |
| Alcohol Consumption | 60269 | -0.028 (-0.054, -0.001) | 0.041 | 74180 | -0.049 (-0.070, -0.029) | 4.05e-06 |
| Binge Drinking | 59677 | 0.013 (-0.019, 0.045) | 0.415 | 73328 | -0.032 (-0.057, -0.006) | 0.01 |
| Caffeine consumption | 55549 | 0.008 (-0.017, 0.033) | 0.536 | 68843 | -0.005 (-0.025, 0.015) | 0.61 |
| BMI | 61794 | 0.061 ( 0.055, 0.068) | 2.04e-73 | 76551 | 0.052 (0.047, 0.057) | 5.19e-85 |
| Smoking Initiation | 62567 | 0.095 ( 0.038, 0.151) | 9.87e-04 | 77146 | 0.038 (-0.007, 0.083) | 0.10 |
| CPD | 10223 | 0.029 ( 0.016, 0.042) | 1.05e-05 | 14628 | 0.008 (-0.001, 0.016) | 0.08 |

Note. All estimates are adjusted for birth year and educational attainment. Non-planners were given the median time to conception (2 months). Results are for both genotyped and non-genotyped individuals.

**Supplementary Table S14. Sensitivity analysis in mothers including non-planners in time to conception Mendelian randomisation analyses compared with planners only**

|  | **Planners only** | | | **Non-planners included** | | |
| --- | --- | --- | --- | --- | --- | --- |
| **Exposure** | **N** | **Beta (95% CI)** | **P-value** | **N** | **Beta (95% CI)** | **P-value** |
| Alcohol consumption | 20,974 | 0.652 (-0.807, 2.111) | 0.381 | 25,079 | -2.653 (-4.193, -1.112) | 0.0007 |
| Caffeine consumption | 19,057 | -0.214 (-1.284, 0.856) | 0.695 | 22,819 | -0.406 (-0.406, -1.423) | 0.43 |
| BMI | 21,344 | 0.028 (-0.015, 0.070) | 0.205 | 25,520 | 0.969 (0.811, 1.129) | 6.06 x 10^-33^ |
| Smoking Initiation | 21,589 | -0.412 (-1.281, 0.458) | 0.354 | 25,814 | 0.330 (-0.060, 0.720) | 0.10 |
| CPD | 3,390 | 0.034 (-0.165, 0.234) | 0.738 | 4,580 | 1.009 (0.147, 1.872) | 0.02 |

Note. Individual-level MR results in the mothers. Betas can be interpreted as risk difference.

**Supplementary Table S15. Frequency of sexual intercourse in the four weeks prior to conception.**

|  | Mothers | | |  |
| --- | --- | --- | --- | --- |
|  | Full Sample |  | Genotyped Sample | |
|  | **N** | **Mean (SD)/%** | **N** | **Mean (SD)/%** |
| *Never* | 541 | 0.65% | 132 | 0.48% |
| *Less than 1-2 times every 2 weeks* | 3,412 | 4.09% | 1061 | 3.88% |
| *1-2 times every two weeks* | 11,452 | 13.72% | 3,648 | 13.33% |
| *1-2 times a week* | 29,920 | 35.84% | 9,841 | 35.95% |
| *3-4 times a week* | 27,839 | 33.35% | 9,383 | 34.28% |
| *5-6 times a week* | 6,812 | 8.16% | 2,258 | 8.25% |
| *Every day* | 3,506 | 4.20% | 1,048 | 3.83% |
|  | **Fathers** |  |  |  |
|  | Full Sample |  | Genotyped Sample | |
|  | **N** | **Mean (SD)/%** | **N** | **Mean (SD)/%** |
| *Never* | 348 | 0.52% | 115 | 0.44% |
| *Less than 1-2 times every 2 weeks* | 2,526 | 3.75% | 991 | 3.79% |
| *1-2 times every two weeks* | 9,094 | 13.50% | 3,523 | 13.48% |
| *1-2 times a week* | 24,249 | 36.00% | 9,483 | 36.27% |
| *3-4 times a week* | 22,852 | 34.93% | 8,960 | 34.27% |
| *5-6 times a week* | 5,510 | 8.18% | 2,083 | 7.97% |
| *Every day* | 2,774 | 4.12% | 987 | 3.78% |

**Supplementary Table S16. Evidence for heterogeneity: Cochran’s Q statistics**

| Exposure | Outcome | Q | df | P Value |
| --- | --- | --- | --- | --- |
| BMI | Age at First Birth | 268.13 | 94 | 1.18 x 10^-18^ |
|  | Parity | 200.16 | 90 | 2.35 x 10^-10^ |
|  | Number of miscarriages | 121.15 | 94 | 0.03 |
| Alcohol Frequency | Age at First Birth | 254.81 | 93 | 5.44 x 10^-17^ |
|  | Parity | 224.34 | 82 | 3.60 x 10^-15^ |
|  | Number of miscarriages | 128.72 | 93 | 0.008 |
| Smoking Initiation | Age at First Birth | 1052.53 | 321 | 1.16 x -78 |
|  | Parity | 871.59 | 322 | 3.52 x 10^-52^ |
|  | Number of miscarriages | 356.24 | 321 | 0.09 |
| Caffeine Consumption | Age at First Birth | 12.25 | 5 | 0.03 |
|  | Parity | 2.74 | 5 | 0.74 |
|  | Number of miscarriages | 1.87 | 5 | 0.87 |

**Supplementary Table S17. The MR Egger intercept test: Evidence for bias from horizontal pleiotropy**

| Exposure | Outcome | Intercept (95% CI) | P Value |
| --- | --- | --- | --- |
| BMI | Age at first birth | -0.001 (-0.005, 0.003) | 0.65 |
|  | Parity | -0.0003 (-0.003, 0.003) | 0.83 |
|  | Number of miscarriages | -0.0004 (-0.003, 0.002) | 0.74 |
| Alcohol Frequency | Age at first birth | -0.002 (-0.004, 0) | 0.07 |
|  | Parity | 0.0007 (-0.001, 0.003) | 0.49 |
|  | Number of miscarriages | -0.0002 (-0.002, 0.001) | 0.78 |
| Smoking Initiation | Age at first birth | -0.001 (-0.003, 0.001) | 0.15 |
|  | Parity | 0.001 (-0.0003, 0.003) | 0.11 |
|  | Number of miscarriages | -0.0002 (-0.001, 0.001) | 0.79 |
| Caffeine Consumption | Age at first birth | -0.02 (-0.036, -0.004) | 0.07 |
|  | Parity | 0.0001 (-0.012, 0.013) | 0.98 |
|  | Number of miscarriages | 0.006 (-0.01, 0.021) | 0.52 |

**Supplementary Table S18. Steiger filtering test for possible reverse causation**

| Exposure | Outcome | N SNPs | N After Steiger Filtering | % True |
| --- | --- | --- | --- | --- |
| BMI | Age at First Birth | 96 | 96 | 100 |
|  | Parity | 91 | 91 | 100 |
|  | Number of miscarriages | 96 | 96 | 100 |
| Alcohol Frequency | Age at First Birth | 96 | 80 | 83 |
|  | Parity | 83 | 83 | 100 |
|  | Number of miscarriages | 96 | 87 | 91 |
| Smoking Initiation | Age at First Birth | 368 | 267 | 73 |
|  | Parity | 323 | 278 | 86 |
|  | Number of miscarriages | 368 | 352 | 96 |
| Caffeine Consumption | Age at First Birth | 6 | 6 | 100 |
|  | Parity | 6 | 6 | 100 |
|  | Number of miscarriages | 6 | 6 | 100 |

**Supplementary Table S19. Test of instrument strength and the suitability of the instrument for MR Egger**

| Exposure | Outcome | mF | Unweighted I^2^_GX_ | Weighted I^2^_GX_ |
| --- | --- | --- | --- | --- |
| BMI | Age at First Birth | 59.87 | 0.904 | 0.889 |
|  | Parity | 59.87 | 0.904 | 0.889 |
|  | Number of miscarriages | 59.87 | 0.904 | 0.889 |
| Alcohol Frequency | Age at First Birth | 14.87 | 0.827 | 0.790 |
|  | Parity | 14.72 | 0.825 | 0.832 |
|  | Number of miscarriages | 14.87 | 0.827 | 0.792 |
| Smoking Initiation | Age at First Birth | 7.72 | 0.08 | 0 |
|  | Parity | 7.70 | 0.077 | 0.046 |
|  | Number of miscarriages | 7.72 | 0.08 | 0 |
| Caffeine Consumption | Age at First Birth | 27.83 | 0.268 | 0.172 |
|  | Parity | 27.83 | 0.268 | 0.177 |
|  | Number of miscarriages | 27.83 | 0.268 | 0.175 |

**Supplementary Table S20. Exploratory multivariable Mendelian randomisation estimating the direct effects of smoking initiation and BMI on age at first birth, accounting for education and impulsivity**

| Exposure |  | mF | N SNP | Beta (95% CI) | P Value |
| --- | --- | --- | --- | --- | --- |
| Educational attainment^a^ | Univariable MR | 88.67 | 67 | 0.595 (0.518, 0.672) | 2.40 x 10^-51^ |
|  | Multivariable MR accounting for SI | 5.28 | 182 | 0.621 (0.528, 0.714) | <2.2 x 10^-16^ |
|  | Multivariable MR accounting for BMI | 11.91 | 88 | 0.639 (0.514, 0.764) | 3.59 x 10^-16^ |
| ADHD^b^ | Univariable MR | 13.67 | 9 | -0.108 (-0.150, -0.067) | 2.69 x 10^-7^ |
|  | Multivariable MR accounting for SI | 2.36 | 180 | -0.096 (-0.133, -0.059) | 1.74 x 10^-6^ |
|  | Multivariable MR accounting for BMI | 4.82 | 78 | -0.109 (-0.147, -0.071) | 2.67 x 10^-7^ |
| Smoking initiation | Univariable MR | 7.70 | 352 | -0.272 (-0.31, -0.234) | 3.45 x 10^-45^ |
|  | Multivariable MR accounting for ADHD | 3.09 | 180 | -0.435 (-0.591, -0.279) | 1.70 x 10^-7^ |
|  | Multivariable MR accounting for educational attainment | 4.23 | 182 | -0.403 (-0.527, -0.279) | 1.43 x 10^-9^ |
| Body Mass Index | Univariable MR | 4.76 | 94 | -0.112 (-0.163, -0.061) | 1.66 x 10^-5^ |
|  | Multivariable MR accounting for ADHD | 24.19 | 78 | -0.513 (-0.106, 0.003) | 0.07 |
|  | Multivariable MR accounting for educational attainment | 29.65 | 88 | -0.056 (-0.113, 0.0008) | 0.06 |

Note. a = educational attainment was measured as years in education. There were 74 independent genome-wide significant SNPs (Okbay et al., 2016).

b = attention deficit hyperactivity disorder (ADHD) was used as a proxy for impulsivity (Demontis et al., 2019). There were 9 SNPs independently associated with ADHD at p<5x10-6.

**References**

Demontis, D., Walters, R. K., Martin, J., Mattheisen, M., Als, T. D., Agerbo, E., Baldursson, G., Belliveau, R., Bybjerg-Grauholm, J., Bækvad-Hansen, M., Cerrato, F., Chambert, K., Churchhouse, C., Dumont, A., Eriksson, N., Gandal, M., Goldstein, J. I., Grasby, K. L., Grove, J., … Neale, B. M. (2019). Discovery of the first genome-wide significant risk loci for attention deficit/hyperactivity disorder. *Nature Genetics*, *51*(1), 63–75. https://doi.org/10.1038/s41588-018-0269-7

Ducci, F., Kaakinen, M., Pouta, A., Hartikainen, A.-L., Veijola, J., Isohanni, M., Charoen, P., Coin, L., Hoggart, C., Ekelund, J., Peltonen, L., Freimer, N., Elliott, P., Schumann, G., & Järvelin, M.-R. (2011). TTC12-ANKK1-DRD2 and CHRNA5-CHRNA3-CHRNB4 influence different pathways leading to smoking behavior from adolescence to mid-adulthood. *Biological Psychiatry*, *69*(7), 650–660. https://doi.org/10.1016/j.biopsych.2010.09.055

Fowler, C. D., Lu, Q., Johnson, P. M., Marks, M. J., & Kenny, P. J. (2011). Habenular α5* nicotinic receptor signaling controls nicotine intake. *Nature*, *471*(7340), 597–601. https://doi.org/10.1038/nature09797

Helgeland, Ø., Vaudel, M., Sole-Navais, P., Flatley, C., Juodakis, J., Bacelis, J., Koløen, I. L., Knudsen, G. P., Johansson, B. B., Magnus, P., Kjennerud, T. R., Juliusson, P. B., Stoltenberg, C., Holmen, O. L., Andreassen, O. A., Jacobsson, B., Njølstad, P. R., & Johansson, S. (2021). Characterization of the genetic architecture of BMI in infancy and early childhood reveals age-specific effects and implicates pathways involved in Mendelian obesity. *MedRxiv*, 2021.05.04.21256508. https://doi.org/10.1101/2021.05.04.21256508

Manichaikul, A., Mychaleckyj, J. C., Rich, S. S., Daly, K., Sale, M., & Chen, W.-M. (2010). Robust relationship inference in genome-wide association studies. *Bioinformatics*, *26*(22), 2867–2873. https://doi.org/10.1093/bioinformatics/btq559

Millard, L. A. C., Munafò, M. R., Tilling, K., Wootton, R. E., & Smith, G. D. (2019). MR-pheWAS with stratification and interaction: Searching for the causal effects of smoking heaviness identified an effect on facial aging. *PLOS Genetics*, *15*(10), e1008353. https://doi.org/10.1371/journal.pgen.1008353

Munafò, M. R., Timofeeva, M. N., Morris, R. W., Prieto-Merino, D., Sattar, N., Brennan, P., Johnstone, E. C., Relton, C., Johnson, P. C. D., Walther, D., Whincup, P. H., Casas, J. P., Uhl, G. R., Vineis, P., Padmanabhan, S., Jefferis, B. J., Amuzu, A., Riboli, E., Upton, M. N., … Davey Smith, G. (2012). Association Between Genetic Variants on Chromosome 15q25 Locus and Objective Measures of Tobacco Exposure. *JNCI: Journal of the National Cancer Institute*, *104*(10), 740–748. https://doi.org/10.1093/jnci/djs191

Okbay, A., Beauchamp, J. P., Fontana, M. A., Lee, J. J., Pers, T. H., Rietveld, C. A., Turley, P., Chen, G.-B., Emilsson, V., Meddens, S. F. W., & others. (2016). Genome-wide association study identifies 74 loci associated with educational attainment. *Nature*, *533*(7604), 539–542.

Thorgeirsson, T. E., Geller, F., Sulem, P., Rafnar, T., Wiste, A., Magnusson, K. P., Manolescu, A., Thorleifsson, G., Stefansson, H., Ingason, A., Stacey, S. N., Bergthorsson, J. T., Thorlacius, S., Gudmundsson, J., Jonsson, T., Jakobsdottir, M., Saemundsdottir, J., Olafsdottir, O., Gudmundsson, L. J., … Stefansson, K. (2008). A Variant Associated with Nicotine Dependence, Lung Cancer and Peripheral Arterial Disease. *Nature*, *452*(7187), 638–642. https://doi.org/10.1038/nature06846

Tobacco Consortium. (2010). Genome-wide meta-analyses identify multiple loci associated with smoking behavior. *Nature Genetics*, *42*(5), 441–447.
